# Optimisation of and mechanisms behind neural distortion responses obtained with cochlear-implant stimulation by interleaved pulse trains

**DOI:** 10.64898/2026.11.09.26367183

**Authors:** Charlotte Garcia, François Guérit, Dorothée Arzounian, Robert P. Carlyon

## Abstract

**Purpose:** We investigated the mechanisms behind the generation of Neural Distortion Responses (NDRs) measured with electroencephalography when interleaving amplitude-modulated (AM) pulse trains in cochlear implant (CI) users.

**Methods:** Previous experiments measured NDRs using interleaved pulse trains in which odd-and even-numbered pulses were respectively modulated at F1 and F2 Hz, where F1=2*F2/3 and F1≈80Hz. Experiment 1 separated quadratic and cubic distortion products using a relationship of F1 and F2 such that F1 = (2*F2/3)+2. Experiment 2 retained the original F1=2*F2/3 relationship but shifted the starting AM phase of F2 relative to that of F1. Experiment 3 simultaneously separated the quadratic and cubic distortion products and varied the relative phase of F1 and F2. Experiment 4 flipped the leading polarity of the second pulse in each pulse-pair such that the central phase of the resulting pseudo-triphasic pulses was anodic. All experiments were conducted with Cochlear CI users and interpreted using two computational models of the electrically stimulated auditory nerve.

**Results:** Both quadratic (QDR) and cubic distortion products (CDR) were observed. Varying relative AM phase had an effect on NDR amplitudes, with the largest amplitude when maxima of the 2 AM patterns were aligned. Stimulating with an anodic central phase within the AM pulse pairs resulted in stronger responses for 4 of 9 CI users. Computational modelling suggested a stronger role for refractoriness than for facilitation in generating the NDR, but failed to fully account for the effect of flipping the polarity.

**Conclusion:** NDRs consist of both quadratic and cubic components, and are greatest when maxima of the F1 and F2 pulse trains align. Refractoriness is likely more important than facilitation for generating the NDR.

## Introduction

Cochlear implants (CIs) are neuro-prosthetic devices that have provided a sense of sound to over one million users worldwide by directly stimulating the auditory nerve [1]. Although many listeners achieve improved speech-perception using their CI there is a lot of variability between users [2]. This variability has inspired numerous research studies investigating its causes and ways in which CI stimulation can be optimised for improved speech-perception outcomes [3], [4]. One approach is development of fitting procedures that leverage objective electrophysiological measures reflective of perceptual reports of loudness, with the aim of optimising the stimulation current levels of the CI. This has been tried primarily using evoked potential recordings from the auditory nerve (electrically evoked compound action potentials, or ECAPs) and brainstem (electrically evoked auditory brainstem responses, or EABRs), with limited and variable correlations with loudness perception [5], [6], [7]. A key limitation of these approaches is that they usually present electrical-current stimulation pulses at a slow rate (< 80 pulses per second, or pps) and measure the responses in the gaps between the stimuli [8]. These slow rates are necessary in part to extract the neural response from contamination by the large electrical artefact produced by CI stimulation, but they render the stimulation very different to that used in CI users’ every-day ‘coding strategies.’ It is therefore desirable to have a technique for measuring neural responses to electrical stimulation in CIs that is as close as possible to those leveraged in everyday use and that isolates these responses from the ubiquitous electrical artefacts.

Carlyon *et al* proposed the ALternating Frequency Interleaved Electrical Stimulation (ALFIES) technique as a way to measure the sustained neural response of CI users to high-rate, amplitude-modulated (AM) pulse trains presented to one or two electrodes in an interleaved manner [9]. The high rate and the amplitude modulation both render the stimulus more similar to stimulation CI users listen to every day, compared to the stimuli used for ECAPs and EABRs. In ALFIES, two interleaved pulse trains are amplitude-modulated at different frequencies F1 and F2 Hz. When F1 and F2 were 80 and 120 Hz, respectively, electroencephalography (EEG) recordings revealed the presence of a neural distortion response (NDR) at 40 Hz. This component is unlikely to reflect nonlinearities in the CI stimulation system because the two AM pulse trains are interleaved in time and nonlinearities from the device occur almost instantaneously. Conversely, the auditory system responds to each pulse in the stimulation pattern in a way that is not independent of adjacent pulses, via mechanisms that include refractoriness and facilitation at the level of the auditory nerve [10]. This “undoes” the independence of the F1 and F2 pulse trains, allowing subsequent and/or simultaneous nonlinearities within the auditory system to produce NDRs at composite frequencies such as F2-F1. The NDR had a group delay of approximately 45 milliseconds (ms), consistent with a thalamic and/or cortical neural generator and inconsistent with the zero delay expected from an electrical artefact. Furthermore, Guérit *et al* suggested that the temporal integration mechanism that precedes neural nonlinearities to generate an NDR likely occurs as early as the auditory nerve membrane and cochlear nucleus, with these NDRs being propagated to cortical/thalamic generators where they can be detected using EEG [11]. This was evidenced by the decrease in NDR amplitude as the inter-pulse interval (IPI) between F1 and F2 pulses was increased above 200-400 μs, with no measurable response > 900μs [11]. Thalamic and cortical integration mechanisms would be expected to have longer time constants.

The experiments described here had two broad goals. The first was to improve our understanding of the neural processes revealed by the ALFIES method. As noted above, the presence of the NDR demonstrates the operation of some between-pulse temporal dependency or dependencies, followed by one or more nonlinearities. Understanding the temporal dependencies between pulses is of particular interest given the trend of increasing CI stimulation rates, and hence shorter inter-pulse intervals, in modern CI processing strategies. Although Guérit *et al* showed that the dependencies responsible for the NDR had a short time constant reflective of a peripheral basis [11], those data do not distinguish between specific mechanisms. We therefore use two computational models that, respectively, incorporate facilitation and refractoriness, and evaluate their predictions of the effect of the relative phase of the F1 and F2 modulations on the NDR. Our experimental results suggest that refractoriness plays the greater role. We also build on previous ALFIES studies to investigate the nature of the nonlinearity responsible for NDR generation. The NDR investigated previously could have reflected either a quadratic (F2-F1) and/or a cubic (2*F1-F2) distortion component, as both give rise to distortions at the same frequency when F2 = 1.5*F1. While these components were intentionally confounded for the purpose of maximising potential SNR, it was not known whether the NDR is quadratic in nature, cubic, or a mixture of both. In our first experiment, we separated the quadratic (QDR) and cubic distortion responses (CDR) by changing F1 to 82 Hz while keeping F2 at 120 Hz, resulting in separated quadratic and cubic distortion products at 38 and 44 Hz, respectively. We found both QDRs and CDRs with group delays consistent with thalamo-cortical neural generators. Given this new evidence that both distortions contribute to the NDR, the importance of optimising parameters such that they interfere constructively became clear, both to maximise SNR and to achieve the most faithful representation of the response of the auditory system.

Experiments 2 to 4 addressed our second motivation, which was to maximise the size of the NDR and to refine the use of ALFIES as a clinical technique. ALFIES already shows promise in this regard: it measures the sustained response to high-rate pulse-trains while avoiding electrical artefact, requires only a single pair of recording electrodes and standard EEG equipment, has a steep amplitude growth function, and – by presenting the F1 and F2 pulse trains to electrodes separated by varying amounts – has the potential to measure the selectivity as well as sensitivity to CI stimulation [9], [11]. A potential limitation is that the amplitude of the NDR reported in previous studies was lower than that of the electrically evoked steady-state response (eASSR). We show here that by presenting both the F1 and F2 modulations such that their maxima are aligned, it is possible to produce an NDR amplitude that is comparable to that produced by the eASSR, thereby combining the benefits of the two techniques. Experiment 4 also shows that the NDR may be further increased for some participants by presenting the F1 and F2 pulses in opposite leading polarity.

## Methods

### Research Participants

A total of 16 CI users implanted with devices from Cochlear Ltd (Sydney, Australia) took part (Table 1). Two were bilaterally implanted and participated in the first experiment with both ears, leading to a total of 18 ears tested. Ethical approval was obtained for the experiments from the National Research Ethics Committee for the East of England (ref. 118230), and written informed consent was collected prior to participation. Data was collected between the years of 2023 and 2025 at the MRC Cognition & Brain Sciences Unit at the University of Cambridge, U.K. Each participant is anonymised and identified by an ID (see Table 1) from which they cannot be re-identified. The participants’ associated IDs are only known to researchers within the Cambridge Hearing Group at the University of Cambridge.

**Table 1:**
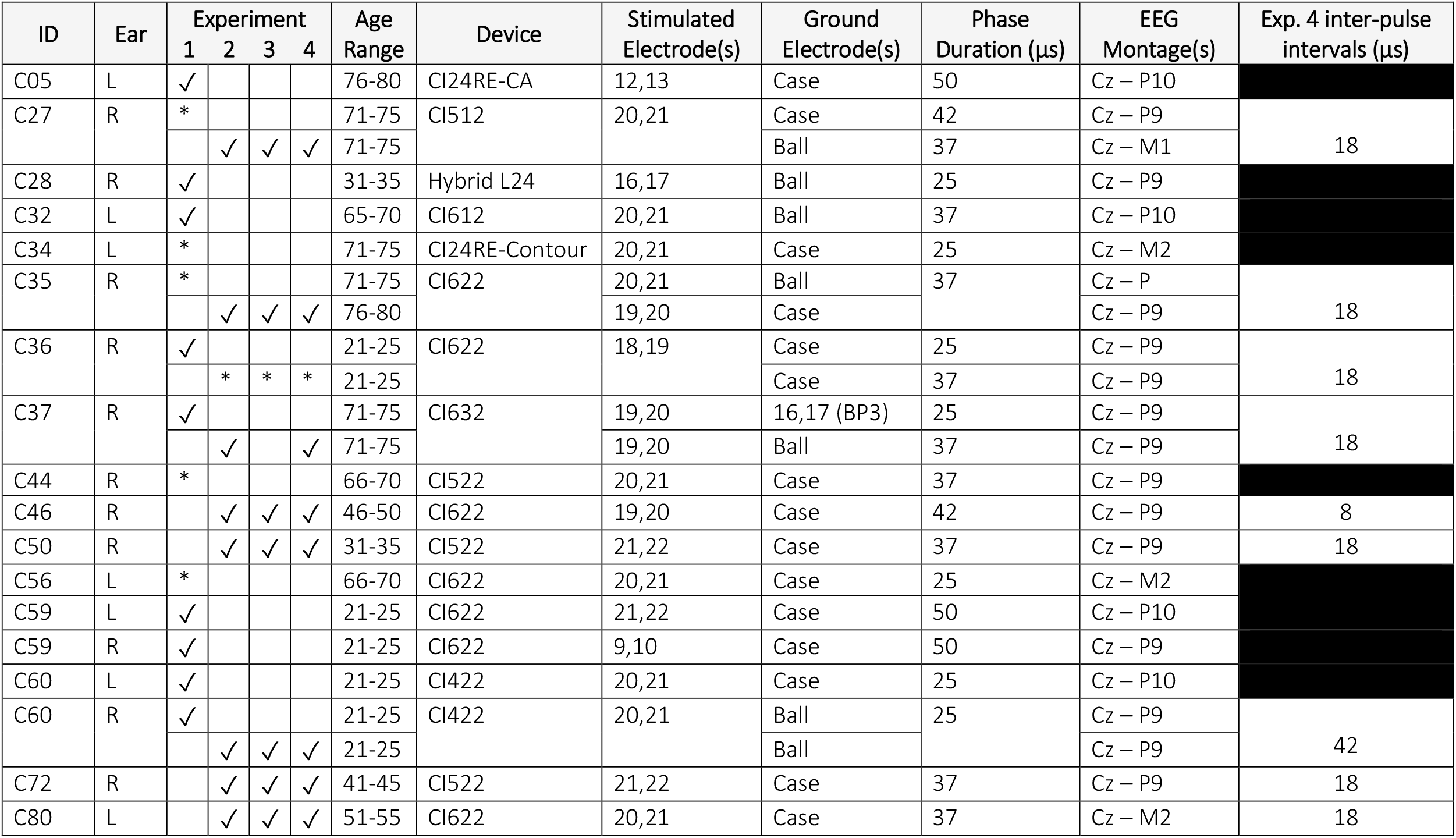
Participant Information (* indicates exclusion from analysis due to lack of significance of distortion products or group delays from 0 ms)

| ID | Ear | Experiment |  |  |  | Age Range | Device | Stimulated Electrode(s) | Ground Electrode(s) | Phase Duration (µs) | EEG Montage(s) | Exp. 4 inter-pulse intervals (µs) |
|---|---|---|---|---|---|---|---|---|---|---|---|---|
|  |  | 1 | 2 | 3 | 4 |  |  |  |  |  |  |  |
| C05 | L | ✓ |  |  |  | 76-80 | CI24RE-CA | 12,13 | Case | 50 | Cz – P10 |  |
| C27 | R | * |  |  |  | 71-75 | CI512 | 20,21 | Case | 42 | Cz – P9 | 18 |
|  |  |  | ✓ | ✓ | ✓ | 71-75 |  |  | Ball | 37 | Cz – M1 |  |
| C28 | R | ✓ |  |  |  | 31-35 | Hybrid L24 | 16,17 | Ball | 25 | Cz – P9 |  |
| C32 | L | ✓ |  |  |  | 65-70 | CI612 | 20,21 | Ball | 37 | Cz – P10 |  |
| C34 | L | * |  |  |  | 71-75 | CI24RE-Contour | 20,21 | Case | 25 | Cz – M2 |  |
| C35 | R | * |  |  |  | 71-75 | CI622 | 20,21 | Ball | 37 | Cz – P | 18 |
|  |  |  | ✓ | ✓ | ✓ | 76-80 |  | 19,20 | Case |  | Cz – P9 |  |
| C36 | R | ✓ |  |  |  | 21-25 | CI622 | 18,19 | Case | 25 | Cz – P9 | 18 |
|  |  |  | * | * | * | 21-25 |  |  | Case | 37 | Cz – P9 |  |
| C37 | R | ✓ |  |  |  | 71-75 | CI632 | 19,20 | 16,17 (BP3) | 25 | Cz – P9 | 18 |
|  |  |  | ✓ |  | ✓ | 71-75 |  | 19,20 | Ball | 37 | Cz – P9 |  |
| C44 | R | * |  |  |  | 66-70 | CI522 | 20,21 | Case | 37 | Cz – P9 |  |
| C46 | R |  | ✓ | ✓ | ✓ | 46-50 | CI622 | 19,20 | Case | 42 | Cz – P9 | 8 |
| C50 | R |  | ✓ | ✓ | ✓ | 31-35 | CI522 | 21,22 | Case | 37 | Cz – P9 | 18 |
| C56 | L | * |  |  |  | 66-70 | CI622 | 20,21 | Case | 25 | Cz – M2 |  |
| C59 | L | ✓ |  |  |  | 21-25 | CI622 | 21,22 | Case | 50 | Cz – P10 |  |
| C59 | R | ✓ |  |  |  | 21-25 | CI622 | 9,10 | Case | 50 | Cz – P9 |  |
| C60 | L | ✓ |  |  |  | 21-25 | CI422 | 20,21 | Case | 25 | Cz – P10 |  |
| C60 | R | ✓ |  |  |  | 21-25 | CI422 | 20,21 | Ball | 25 | Cz – P9 | 42 |
|  |  |  | ✓ | ✓ | ✓ | 21-25 |  |  | Ball |  | Cz – P9 |  |
| C72 | R |  | ✓ | ✓ | ✓ | 41-45 | CI522 | 21,22 | Case | 37 | Cz – P9 | 18 |
| C80 | L |  | ✓ | ✓ | ✓ | 51-55 | CI622 | 20,21 | Case | 37 | Cz – M2 | 18 |

Some users participated in a single experiment while others participated in three or four. In some users, a significant NDR could not be observed above the noise floor for some experiments, and so they were excluded from further analysis (* in Table 1). Details for significance testing for exclusion are described within each experiment’s section.

### General Experimental Approach

All experiments involved direct stimulation of an adjacent pair of electrodes with interleaved, amplitude-modulated pulse trains using the Nucleus Implant Communicator research software version 4.3 (NIC4.3, nice22.1) provided by Cochlear Ltd. The stimulation parameters were controlled by a laptop using custom software developed in Python version 3.9 (Wilmington, Delaware, USA) that used the NIC4.3 python libraries, and communicated with a CP910 cochlear-implant processor via a programming cable and POD connected to the laptop via USB.

The interleaved AM pulse trains used for stimulation are depicted in Figure 1a-b. Each AM pulse train had a carrier rate of 480 pps, usually with a stimulus onset asynchrony (SOA) of 200 μs (except for experiment 4). These parameters were selected for consistency with previous work and from evidence of no reduction in the NDR amplitude when the AM pulse trains are presented on adjacent electrodes compared to a single electrode, or when the SOA was set to 200-400 μs compared to shorter values [9], [11]. Some experiments (2, 4) used an F1:F2 ratio of 2:3 as in previous work (see Figure 1a), whereas other experiments (1, 3) added 2 Hz to F1 such that the quadratic and cubic distortion product frequencies were isolated but both still fell within ≈ 40 ± 10 Hz, the optimal range for eliciting a cortical response (see Figure 1b; [12], [13]). The precise frequencies used for each experiment are described in each corresponding section.

**Figure 1:**
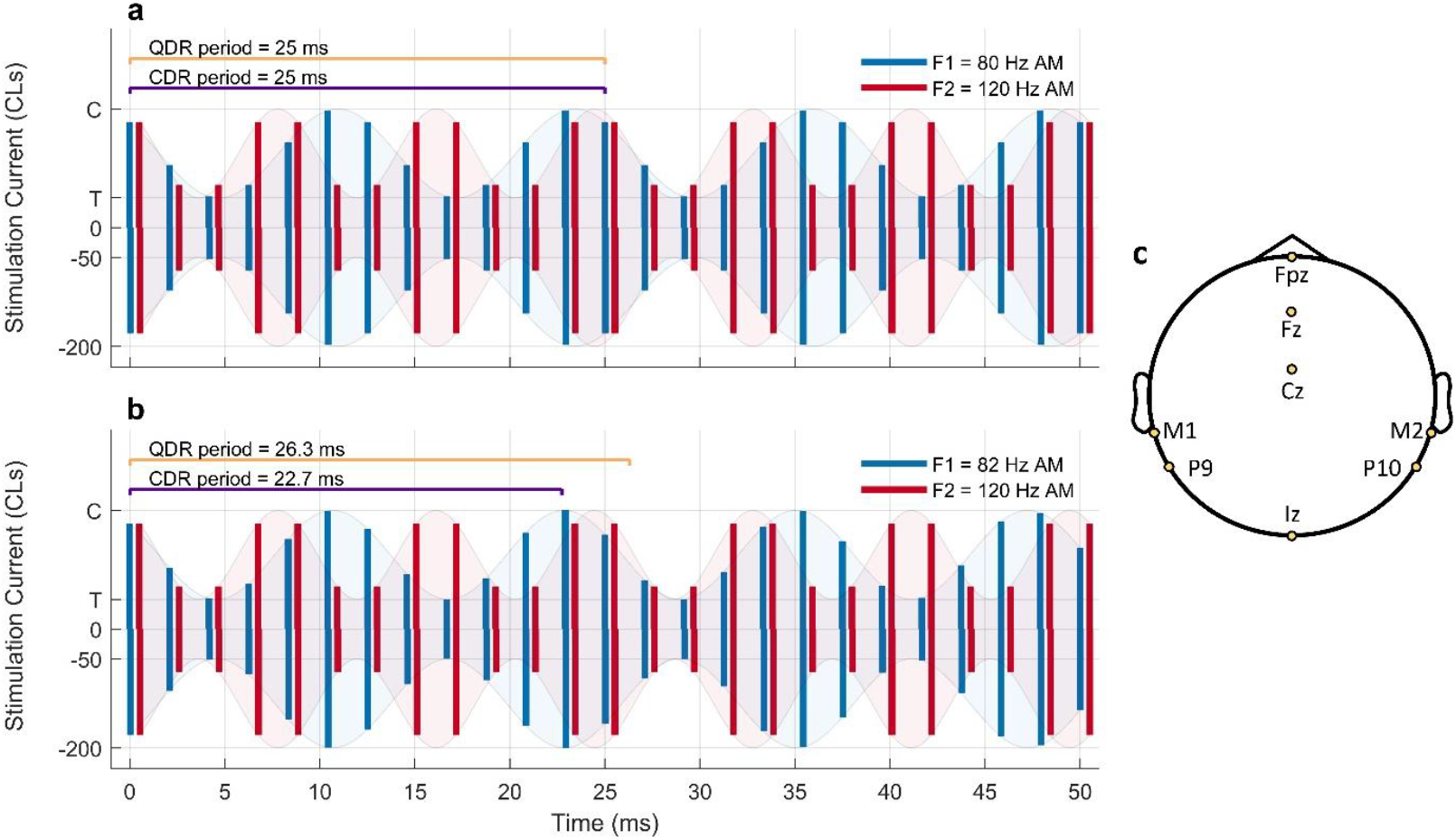
ALFIES stimulation for conditions with 480-pps carrier rates (a) where the quadratic and cubic distortion products (QDR and CDR, respectively) fall on the same frequency, demonstrated here where F1 = 80 Hz (blue) and F2 = 120 Hz (red) resulting in a distortion product frequency of 40 Hz, and (b) where the quadratic and cubic distortion products are separated, demonstrated here where F1 = 82 Hz (blue) and F2 = 120 Hz (red) resulting in a quadratic distortion product of 38 Hz (period displayed in yellow) and a cubic distortion product of 44 Hz (period displayed in purple). (c) EEG montage schematic for the custom 262,144 Hz BioSemi system.

Each stimulating pulse defaulted to a cathodic-leading, biphasic stimulating pulse with an 8-μs inter-phase gap (IPG), unless otherwise indicated. The phase durations and stimulating electrodes differed somewhat across participants (Table 1), with a preference for more-apical electrodes and phase durations that matched those used in the participants’ clinical program. Longer phase durations were used when compliance limits were reached without achieving a loudness rating of “most-comfortable level” (MCL). More-basal electrodes were employed when apical electrodes were switched off in the CI user’s clinical programme or showed atypically high impedance values.

Prior to commencing, impedances were recorded and the upper limit of compliance was calculated for stimulating electrodes. A loudness-scaling procedure was then employed using an 11-point chart where 0 = ‘inaudible’ and 10 = ‘too loud.’ Starting with 0.5-second duration, unmodulated pairs of pulse trains on one electrode that were presented below threshold with slowly increasing current level (with steps of 6 Current Levels or CLs, where 6 CL ≈ 1 dB) until the sounds were just audible. This level was set as the threshold (T), and then the two interleaved trains were amplitude modulated respectively at 80 and 120 Hz between T and a higher current level that was increased in steps of 6 CLs until level ‘4’ (comfortable but too soft) was achieved, steps of 4 CLs until level ‘5’ (comfortable but soft) was achieved, and steps of 2 CLs until level ‘7’ (loud but comfortable) was achieved. The upper modulation current level was then decreased in steps of 1 or 2 CLs to find the highest current level that was considered ‘6’ (most comfortable), and this current level was set as MCL for that electrode. The same procedure was then repeated with the second, adjacent electrode. These two sets of T and MCL levels were then used as the minimum and maximum current between which the pulse trains were amplitude modulated for the interleaved stimulation. The participant was played a 5-10 second segment of the interleaved stimulus to confirm that the level was comfortable to listen to for an extended period of time, and the MCL levels were decreased in steps of 2 CLs if necessary to accommodate this.

Research participants were then asked to remove the research processor, and were fit with an electroencephalography (EEG) cap fitted with 10 contacts (including 2 ground electrodes), placed as depicted in Figure 1c. By default, montages of Cz vs P9 or P10 were used for participants with implants on the Right and Left sides, respectively, leveraging the contact behind the ear contralateral to the stimulating CI. Montages were sometimes adjusted, replacing P9 or P10 with the neighbouring mastoid contact (M1 and M2, respectively), if the default montage provided noisy data with many rejected epochs (see Table 1). The EEG system was custom built by BioSemi B.V. (Amsterdam, Netherlands) with a 262,144 kHz sampling rate. Stimuli were delivered to the CI via direct stimulation using a CP910 processor in the manner described above, in segments of 31 seconds. A trigger was included in the stimulation code at each 1-second interval (excluding the first second to avoid onset responses) throughout stimulation, for use in the analysis pipeline that organised the EEG data into 1-second epochs. The analysis pipeline was employed in real time during data collection as described in Guérit *et al* (2023) and followed the procedure laid out in both Carlyon *et al* (2021) and Guérit *et al* (2023), with some small adjustments described below. It used custom software developed in MATLAB 2020b (Mathworks, Natick, Massachusetts, USA) that reads in and processes EEG data files saved by the ActiView software (ActiHyper version 804) from Biosemi. As a brief summary,this involved averaging the 1-second EEG epochs and converting from the time domain to the frequency domain by calculating a fast Fourier transform (fft) with 1-Hz frequency-bin resolution. Amplitudes in decibels (dB) re 1 μV and phase in radians (rad) were then extracted at frequencies of interest, including both the F1 and F2 AM frequencies (e.g. 80 and 120 Hz, respectively) and the distortion product frequencies (e.g. 40 Hz). A standard procedure was also employed to remove noisy epochs as described in Arzounian *et al* [14], and epochs with noise estimates higher than 2 μV were rejected. A standard of 300 epochs with triggers (5 minutes of steady-state recordings) were recorded for each condition, and real-time analysis of the significance of amplitudes at the NDR frequencies was employed. This standard number of epochs was sometimes extended to 450 or 600 if time allowed in the cases where insignificant responses for the distortion product frequencies were observed after 300 epochs, or reduced to a minimum of 90 epochs in some rare cases if significant and stable responses were observed quickly.

The method for significance testing differed from prior analyses pipelines [9], [11], where significance was confirmed by comparing the amplitude of the target frequency bin in the fft power spectrum to the amplitude in the 3 neighbouring frequencies below and above it. For example, in previous work the amplitude of 40 Hz was compared to the amplitudes of 37 - 39 and 41 - 43 Hz using a criterion F ratio of 10.92 (p < 0.01, approx. 10 dB SNR [15]). Instead, we employed a significance test based on Hotelling T^2^ that compares the amplitude of the target frequency bin to its variation across epochs [13], [16], [17], [18]. This alternative method has the benefit of avoiding the inclusion of (or being close to) 50 Hz in the ‘noise’ frequencies, since some of our frequencies of interest were as high as 47 Hz, and recordings sometimes exhibited the U.K. 50-Hz mains power despite the shielding of the EEG recording booth. Similar to our previous F-ratio approach, the alpha threshold for significance was set to 0.01 instead of the standard 0.05 to reduce the chance of type 1 errors (false positives).

We additionally employed bootstrapping [19] to visualise significance and robustness of individual conditions, as has been used previously for evoked potentials [20] and employed here for steady-state responses in the frequency domain. The analysis pipeline was conducted on subsets of the full-epoch datasets by sampling 1-second epochs with replacement to retain the same total number of epochs. During data collection and for use with real-time analysis, this bootstrapping procedure was only conducted with 10-50 repetitions to allow the computer sufficient time to plot the results in real time. However in post-processing this was done 1000 times. This allowed for calculations of 95% confidence intervals for amplitude and phase values of target frequency bins by ordering the 1000 permutated amplitude and/or phase values in ascending order and identifying the minimum and maximum from the central 950 values. This procedure was also employed for the group delay calculations described below (see experiment 1 methods).

### Modelling Approach

Two phenomenological models of the auditory nerve’s response to electrical stimulation were used. The first model is based on Brochier *et al* (BR22) and includes spike-rate adaptation and refractoriness [21], while the second is based on Joshi *et al* (JO17) and also includes facilitation and accommodation [22]. These served as complementary investigations for each experiment, and were used either to investigate mechanistic explanations for observed results, or to guide parameters for following experiments. This follows work from a preliminary study where we investigated the possible contribution of auditory-nerve refractoriness and/or facilitation to NDR generation at different IPIs [23]. The simulations showed that at short inter-pulse intervals such as used here, NDR generation was primarily driven by facilitation / summation between the F1 and F2 primaries in the JO17 model, while it was primarily driven by refractoriness / masking in the BR22 model.

The BR22 model consists of 1500 neurons distributed along the cochlea. For each combination of stimulating electrode and neuron, a look-up table (re-used from Brochier *et al*.) gives a probability of firing as a function of input current level, based on an inverse cumulative density function of a normal distribution. The 50-% point of each of these functions was derived in that study using single pulses and a combination of a finite-element method conduction model and a cable model of the auditory nerve. The relative spread (~slope) of these functions differs from neuron to neuron and matches group statistics from cat auditory nerve recordings. A stochastic process generates spikes based on such probabilities of firing / look-up table, and if a spike is generated, the input level is elevated to reflect refractoriness and spike-rate adaptation. Absolute refractoriness lasts 700 μs [24], [25], relative refractoriness is modelled by a 6-dB decreased in input current level going back to 0 dB with a time constant of 1.5 ms. We adjusted both the absolute refractory period and relative refractory time constant to 200 μs, as described in Guérit *et al* [23], to better fit the effect of inter-pulse interval on the NDR as reported by Guérit *et al* [11],[26]. This modification is also roughly consistent with recent evidence from ECAP experiments that auditory nerve populations may have absolute refractory periods as short as 300 μs [27]. This however had negligible impact in this study, where all the IPIs where < 200 µs. Adaptation is modelled by two exponential decreases in input stimulation level thresholds with respective time constants of 23 and 212 ms. In the original publication, this was followed by a central integration window and an automatic speech recognition stage, but we ignored these stages here and only analysed the raw “auditory-nerve” spike neurograms.

We created a population-response version of the JO17 model by creating 1500 neurons, matching the single-pulse 50-% firing probabilities of the BR22 model as a function of neuron, stimulating electrode, and level. The original JO17 model consists of two leaky-integrate-and-fire neurons, respectively anodic- and cathodic-sensitive. Time constants of facilitation (~200 µs), refractoriness (600 µs absolute, up to 4-5 ms relative), accommodation and spike-rate adaptation are not parameters per se, but consequences of the time constants of integration, amounts of leakiness and type of noise in the leaky integrate-and-fire process. They match single-fibre cat recordings.

For both models, across all experiments and unless otherwise stated, two seconds of interleaved, amplitude-modulated biphasic pulse-train stimuli (designed to replicate the experimental stimuli) were processed with 60 repetitions. The first second of each segment was ignored to omit any contribution of an onset response and capture only the steady-state output. The modelled neural responses were then cut in 1-second epochs, averaged, summed across all neurons, and the amplitude and phase at the frequencies of interest were extracted with an fft with 1-Hz resolution, as was done for the EEG analysis. Levels were set to recruit approximately 10% of the neurons, which also meant that all modelled NDRs were significantly above the noise floor (assessed with an F test). Both original models are available in open access (BR22: https://gitlab.developers.cam.ac.uk/chg/models/brochier-phenomenological; JO17: https://github.com/neurongeek/ANF-model), as are the modified versions used here [28].

## Experiment 1: quadratic vs cubic nature of the neural distortion

### Experimental Methods

Experiment 1 separated the quadratic and cubic distortion products so as to determine whether both contributed to the observed NDR described by Carlyon *et al* [9] and Guérit *et al* [11], and if so, if they differed in amplitude, phase, and group delay. Twelve CI users took part, two of whom were bilaterally implanted and participated with both ears for a total of fourteen ears (see Table 1).

The nominal-frequency pairs for isolated quadratic (F2-F1) and cubic (2*F1-F2) distortion-product frequencies were 82 & 120 Hz for F1 and F2, respectively, likewise shifted to 76 & 111 Hz and 88 & 129 Hz to calculate the group delays for the QDR (35, 38, and 41 Hz) and CDR (41, 44, and 47 Hz). The AM pulse train carrier rate for each F1 & F2 pair was always set to 4*F2 (444, 480, and 516 Hz), resulting in overall stimulation rates of 888, 920, and 1032 pps, respectively. Amplitude modulation was applied using 0.5*[1 + sin(2π * f_mod_*t + ϕ)], initiated with ϕ = 0π at the start of each 1-second epoch (see Figure 1b), consistent with the AM approach used in previous work presented by Carlyon *et al* and Guérit *et al* [9], [11].

The criteria for inclusion in subsequent analysis was the observation of at least two significant amplitudes at distortion product frequencies (within the QDR *and*/*or* the CDR) at different stimulation rates, in order to enable calculation of a reliable group delay via phase shifts. This excluded participants for whom exactly two significant responses were observed, but only one QDR and one CDR.

Group delays were calculated for the F1 and F2 frequencies as well as the significant QDRs and CDRs according the formula in equation (1):

,
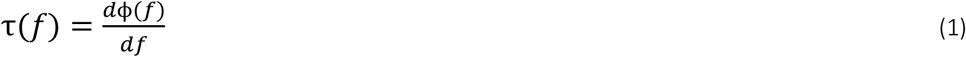

where τ is the group delay in milliseconds (ms), *d*ϕ is the phase shift in radians of the frequency component in question, and *df* is the frequency range in Hz over which the phase shift *d*ϕ occurs. The phases for the neighbouring significant observations were unwrapped prior to group delay calculations when they differed by more than π radians. In the case of three significant responses being observed, *d*ϕ is calculated by fitting a linear regression to the three points. In the case of two significant responses being observed, *d*ϕ is simply defined as the vector connecting the two points, and the *df* is adjusted as needed to reflect the frequency range over which the two significant points are observed. The group delays were calculated for F1, F2, QDR and CDR, and compared to a group delay of 0 ms (as a negative group delay is not physically meaningful) using a one-tailed t-test at the group level, to assess independence from electrical artefacts. The hypothesis was that the group delays for F1 and F2 would not differ significantly from 0, whereas the group delays for QDR and CDR (if present) would be significantly greater than 0 and be consistent with neural generators in the thalamus/auditory cortex. A paired t-test was also performed to assess whether QDRs and CDRs differed from each other.

For each participant for whom significant QDRs and CDRs were both observed for 41 Hz, the amplitudes and phases of these distortion product frequencies were also compared to each other using paired two-tailed t-tests. These statistical tests used a standard α-threshold of 0.05 for 95% confidence, but were further corrected for multiple (7) comparisons, accounting for the seven planned t-tests, to reduce the chance of type 1 errors (false positives), lowering the α-threshold to α = 0.05/7 = 0.0071.

### Modelling Methods

The stimulation conditions used as described above with AM frequency relationships following the rule of F1 = 2*F2/3 + 2 were replicated in both the JO17 and the BR22 model.

### Results

The significance criteria described in the methods section were not achieved for participants C27, C34, C35, C44, and C56 whose data were therefore excluded from analysis. Significant QDRs were observed in all 9 retained ears (from 7 CI users), and significant CDRs were observed in 5 of the 9 retained ears (from 4 CI users).

Two example power spectra from representative participants are displayed in Figure 2. Figure 2a shows data from participant 60R for whom both a QDR (yellow, 41 Hz) and CDR (purple, 47 Hz) were observed. Figure 2d shows data from participant C05L whose data show only a significant QDR (yellow, 35 Hz).

**Figure 2:**
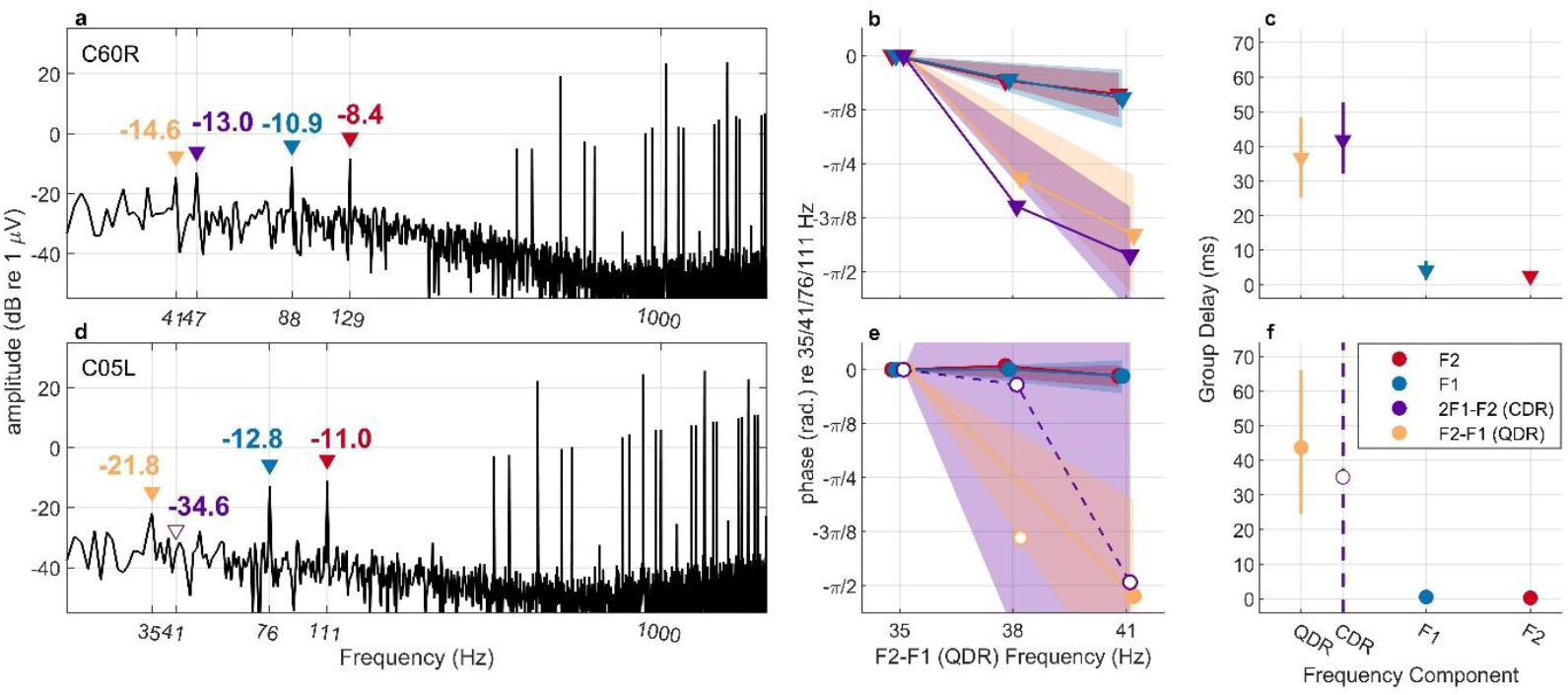
Example power spectra, phase shifts, and group delays for participants C60R (EEG montage: Cz-P9) and C05L (EEG montage: Cz-P10). Throughout: solid symbols and lines represent frequency components with significance from the noise floor after a Hotelling T^2^ analysis procedure, whereas open symbols and dashed lines represent a lack of significance. The color code for the QDR (yellow), CDR (purple), F1 (blue), and F2 (red) frequency components is also consistent throughout. (a) A power spectrum from C60R plotted in black, with the amplitudes of each frequency component of interest QDR (41 Hz), CDR (47 Hz), F1 (88 Hz), F2 (129 Hz) indicated with triangles.(b) Phases for each frequency component from C60R plotted by frequency, with shaded regions representing the central 95% of the distribution of calculated slope values calculated using bootstrapping. (c) Group delays for each frequency component for C60R calculated using equation (1), with error bars representing the central 95% of the distribution calculated using bootstrapping. (d) A power spectrum from C05L, analogous to panel A, where the CDR (41 Hz) is not significantly above the noise floor as represented by the open triangle. (e) Phase shifts for each frequency component from C05L plotted by frequency, with shaded regions representing the central 95% of the distribution of calculated slope values calculated using bootstrapping. It can been seen that for the non-significant CDR, this region extends to positive slopes. (f) Group delays for each frequency component from C05L, analogous to panel C, but showing a non-significant CDR where 95% confidence intervals include 0ms. (EEG = electroencephalography, QDR = quadratic distortion response (F2-F1), CDR = cubic distortion response (2*F1-F2), rad. = radians, α = 0.01)

Group delays were then calculated for each of the four frequencies components of interest (QDR, CDR, F1, and F2) as described above using equation (1) for all significant participant x component combinations that demonstrated significant responses for at least two frequencies, allowing inspection of any phase shifts. Figure 2b shows the phase shifts over frequencies for C60R for each of the four frequency components, and figure 2c shows their associated group delays. Figures 2e and 2f show the analogous results from C05. The 95% confidence intervals were calculated for individual participants’ group delays using the bootstrapping approach described in the Methods section. These intervals encompass zero for the primary components F1 and F2 (as expected for an artefact) but not for the distortion responses with significant amplitudes. Across participants, phase shifts and group delays for each frequency component can be viewed in Figure 3. One-tailed t-tests showed the group delays were significantly above 0 ms for both the QDR and the CDR but not for the primary components F1 and F2 (QDR: t(7) = 6.42, *p* = 0.0002; CDR: t(5) = 17.13, *p* < 0.0001; F1: t(8) =0.35, *p* = 0.37; F2: t(8) = −0.94, *p* = 0.81), as hypothesized. The mean group delay of the QDR was 37.5 ms (95% confidence lower bound = 25.7 ms) and that of the CDR was 36.4 ms (lower bound = 33.0 ms), both consistent with thalamo-cortical responses. The group delay did not differ significantly between the QDR and CDR, as shown in Figure 4a and assessed using a paired-sample two-tailed t test (t(4) = 0.786, *p* = 0.476).

**Figure 3:**
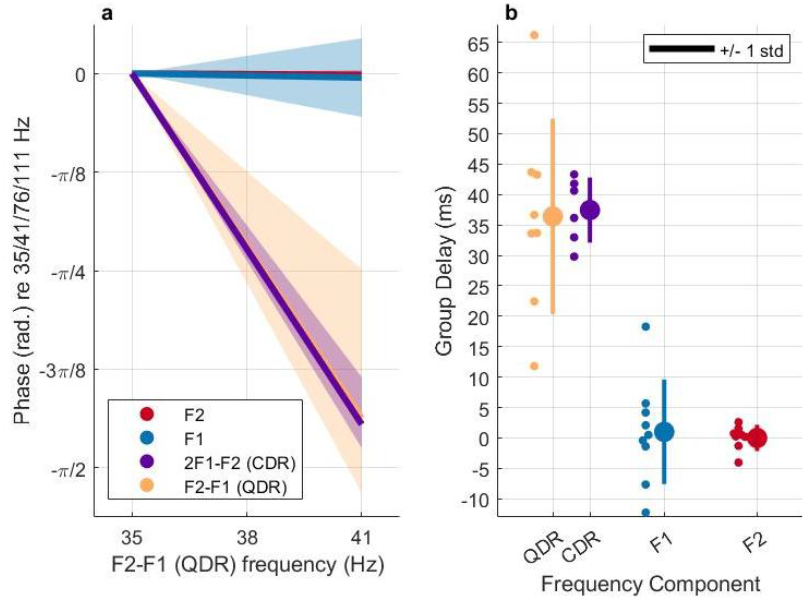
Phase shifts (a) and Group delays (b) for each frequency component. F1 (blue) and F2 (red) show no phase shifts and group delays that are insignificant from 0 ms at the group level, indicating domination by electrical artefact. QDR (yellow) and CDR (purple) both show decreases in phase with increasing frequency, and group delays that are significantly greater than 0 at the group level, with averages around 35-40ms, consistent with a group delay typical of a thalamo-cortical response. Each small dot in (B) represents one participant, and the large dots and error bars indicate group-level averages and ± 1 std. (QDR = quadratic distortion response, CDR = cubic distortion response, std = standard deviation)

**Figure 4:**
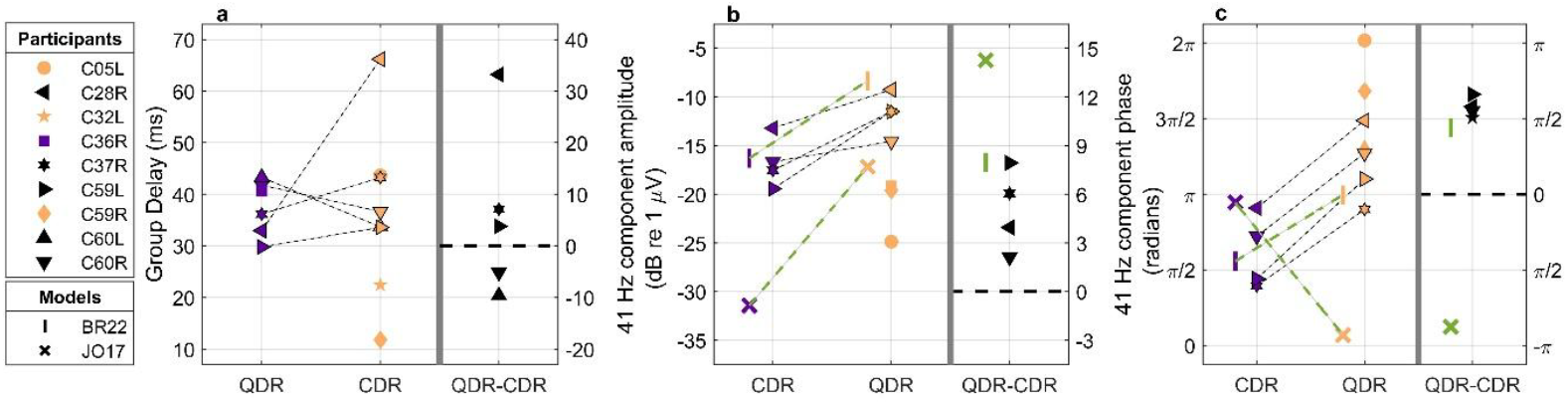
Pairwise comparisons for quadratic and cubic distortion responses for those who had significant responses. Results of the JO17 and BR22 modelling are also included for amplitudes and phases. Throughout, QDRs (purple) and CDRs (yellow) are displayed with reference to the left y-axis, in addition to their pairwise difference (black) with reference to the right y-axis. (a) Group delays can be observed for both QDRs and CDRs for 5 participants, and they do not consistently differ. (b) QDR amplitudes were 4.02 ± 3.02 dB (95% confidence intervals) higher than frequency-matched CDR amplitudes, although this difference did not survive corrections for multiple comparisons. JO17 and BR22 modelling results are indicated using the color green and with crosses and vertical ticks, respectively. (c) A significant and consistent difference averaging 0.57 ± 0.10 π rad. was observed between the phases of the QDRs and CDRs. The legend is color-coordinated such that a purple symbol indicates a participant for whom only a significant CDR was observed at 41 Hz, a yellow symbol represents a participant for whom only a significant QDR was observed at 41 Hz, and a black symbol indicates significance for both QDR and CDR at 41 Hz, and therefore the ability to do a pairwise comparison.

Post-hoc pairwise comparisons were further computed for amplitudes and phases of significant QDR and CDR at a matched frequency (41 Hz) using standard two-tailed t-tests. Only frequency-matched components were compared due to evidence of high variation between CI users for the most-optimal frequency at which cortical steady-state responses are observed, as well as steady-state response amplitudes varying within participants as a function of frequency [13]. The left-hand parts of each panel of Figure 4 show the group delays, amplitudes, and phases of the QDR and CDR for each participant, with the difference between the QDR and CDR shown by the black symbols in the right-hand parts. Significantly higher amplitudes for frequency-matched QDRs were observed compared to CDRs (t(3) = 3.974, *p* = 0.028, Figure 4b), although this comparison does not survive correction for multiple comparisons (α = 0.05/7 = 0.0071). However it should additionally be noted that for three ears (C05, C59R, and C60L), no significant CDR response was observed, whereas QDRs were, which in itself is partially indicative of lower amplitudes for the CDR. Phases of the QDR and CDR were observed to be different from each other (t(3) = −44.009, *p* < 0.0001, see Figure 4c).

The separated distortion products were then modelled using BR22 and JO17 to investigate whether the experimentally observed pattern of amplitudes and phases for the QDR and CDR can be replicated by either model. The same stimulation paradigm applied to the CI users was applied to the models, and the results are shown by the vertical tick (|; BR22) and cross (x; JO17) symbols of Figures 4b and 4c (depicted with green dashed lines in the left-hand panels, and green symbols in the right-hand panels). While both models replicate the lower amplitudes for the frequency-matched CDR than for the QDR (Figure 4b), BR22 appears to match the consistent phase difference between them better than JO17 (Figure 4c).

These results demonstrate the presence of both quadratic and cubic distortion products in response to ALFIES stimulation, and suggest that previous studies observed neural distortion products that were some combination of both quadratic and cubic distortion components. The pairwise comparisons suggest that while in some cases cubic distortion products may not be present or at least below the noise floor of the EEG recordings, they may not be optimally interacting with the quadratic components. This is because the frequency-matched pairwise comparisons between QDR and CDR showed a robust and consistent phase difference of ≈ π/2, compared to the zero difference required for perfect summation.

The modelling work also suggests that the observed QDR and CDR are more dominated by the effects of refractoriness than facilitation. This can be seen in Figs. 4b-c where the BR22 model (where NDR generation is dominated by refractoriness) more closely replicates the experimental data both in terms of the relative strength of the QDR and CDR but importantly, also in terms of their phase relationship.

### Experiment 2: maximising constructive interference; the effects of relative AM phase

The objective of experiment 2 was to determine whether there was an effect of the relative AM phase of the two AM pulse trains on the amplitude of the neural distortion response when the QDR and CDR were confounded (i.e. at the same frequency). If this effect exists, it was also of interest to determine the optimal relative AM phase of the two AM pulse trains to achieve the highest NDR amplitude and therefore also the highest SNR.

In earlier experiments (i.e. Carlyon *et al*, 2021; Guérit *et al*, 2023) the amplitude modulation of the F1 and F2 pulse trains was implemented using *sin()* with starting phases of ϕ=0, resulting in the mid-amplitude portions of the two envelopes being temporally aligned (as in Figure 5b). For simplicity and for Experiment 2 onward, the amplitude modulation was implemented using *cos()* to allow for maxima to be aligned with starting phases of ϕ=0.

**Figure 5:**
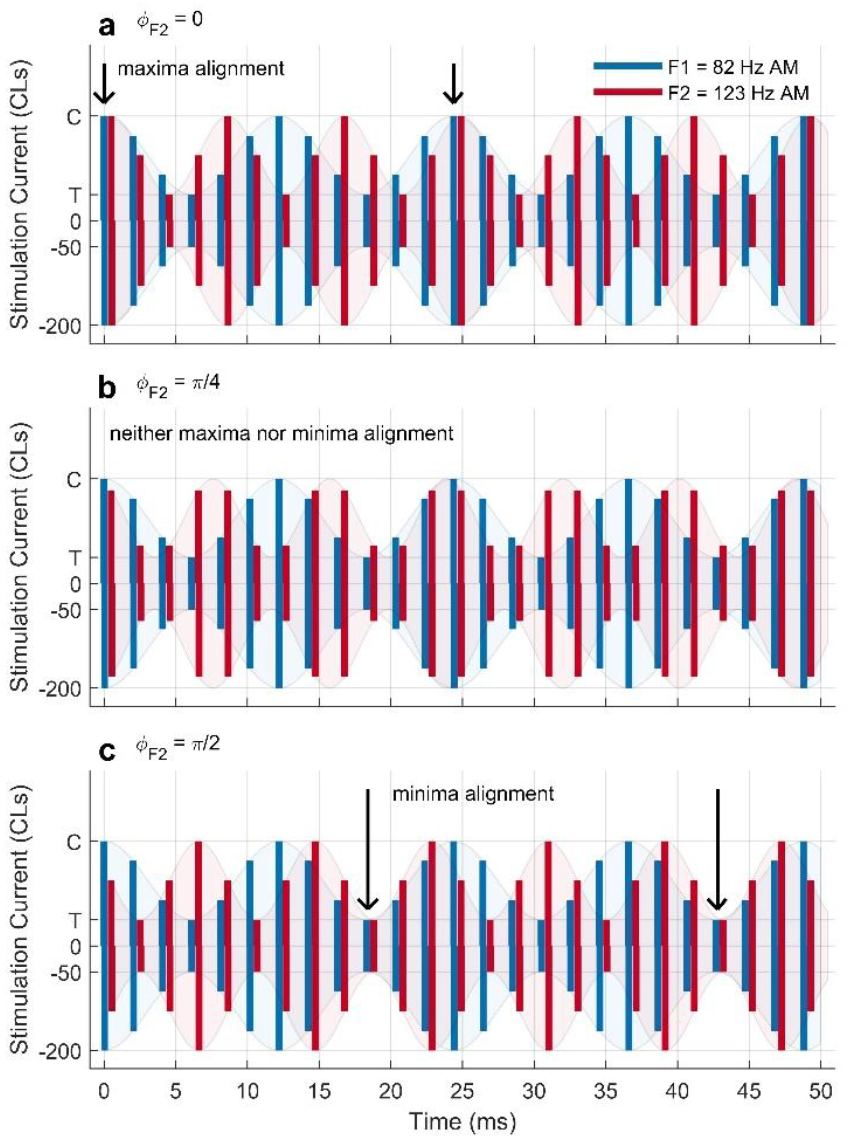
Stimulation conditions for ALFIES interleaved, amplitude modulated pulse trains with varying relative AM phases. F1 (82 Hz, blue pulses) always starts at ϕ_F1_ = 0 radians at t = 0 ms. F2 Hz (123 Hz, red pulses) vary in their starting phases between panels. In panel (a), ϕ_F2_ = 0 radians at t = 0 ms, and it can be observed that the maxima of the F1 and F2 modulated pulse trains are temporally aligned. In panel (b), ϕ_F2_ = π/4 radians at t = 0 ms, and neither the maxima nor the minima are aligned. This represents the amplitude modulation alignment present in previous studies. In panel (c), ϕ_F2_ = π/2 radians at t = 0 ms, and the minima are temporally aligned.

This experiment was guided by modelling predictions done prior to the experimental portion. The second objective was therefore to compare the observed results with the modelling predictions to investigate which temporal dependency (i.e. refractoriness or facilitation) is most likely to undo the interleaving of the stimulus and give rise to the distortion response.

### Modelling Methods

The amplitude modulation of the F1 and F2 pulse trains were set respectively to 82 Hz and 123 Hz, resulting in an NDR at 41 Hz (QDR and CDR superimposed). The F1 pulse train was amplitude modulated using 0.5*[1 + cos(2π *f_mod_* t + ϕ)] initiated with ϕ = 0 at the start of each 1-second epoch, resulting in an initial amplitude at the maximum stimulating current within the amplitude modulation. The F2 pulse train was amplitude modulated with ϕ varied between 0 and 2π at the start of each 1-second epoch in increments of π/16. Figure 5 depicts the stimulation conditions for an F2 starting phase of 0 (a, where the maxima of F1 and F2 are aligned), π/4 (b, consistent with the relative phase used in experiment 1 where neither the maximum nor the minimum current levels are aligned between the two pulse trains), and π/2 (c, where the minima are aligned), respectively. For both models, the amplitude and the phase of the NDR response at 41 Hz was observed as a function of the F2 starting AM phase. Due to stochasticity in the models, each condition was run 60 times for a period of 2 seconds. The 1^st^ second of each was excluded from analysis to avoid inclusion of onset effects, resulting in a total of 60 1-second epochs that were included to calculate the amplitudes and phases of the NDR frequency component in the ffts, as well as their variation.

### Modelling Results

The amplitudes of the ALFIES NDR components according to the JO17 and BR22 models are depicted in Figure 6a as a function of the varying F2 starting phase. Note that the absolute values of the NDR amplitudes and associated modulations within and between models are somewhat arbitrary, as these are determined based on specific model parameters (such as the number of neurons and their threshold distributions), but the overall pattern and position of the fluctuations shown are largely robust to these parameters. The BR22 model yields the highest NDR amplitudes when the peaks of the AM are aligned between the two pulse trains,i.e. at ϕ_F2_ = 0 and ϕ_F2_ = π, and the lowest NDR amplitudes when the troughs are aligned, i.e. at ϕ_F1_ = π/2. A different pattern is observed for the JO17 model, with the highest NDR amplitudes between π/2 and 5π/8 and the lowest amplitudes near 3π/8. For both models the variation in predicted NDR amplitude between 0 and π radians is repeated between π and 2π. Within this 0-π period, the BR22 model’s amplitude predictions oscillate between high and low once, whereas the JO17 model does this twice. As shown in Figure 6b, the models predict phases for the NDRs that show the same increase in phase with increasing F2 starting phase, but are π out of phase with each other. This is likely because – as shown in Guérit *et al* 2025 – masking of the F2 pulses happen in the BR22 model when peaks are aligned due to refractoriness, while summation occurs in JO17 in the same scenario due to facilitation [23]. Therefore, the largest amplitude within a cycle of the modelled NDR response changes depending on which model is used (and therefore on whether facilitation or refractoriness dominates the interaction between F1 and F2 pulses).

**Figure 6:**
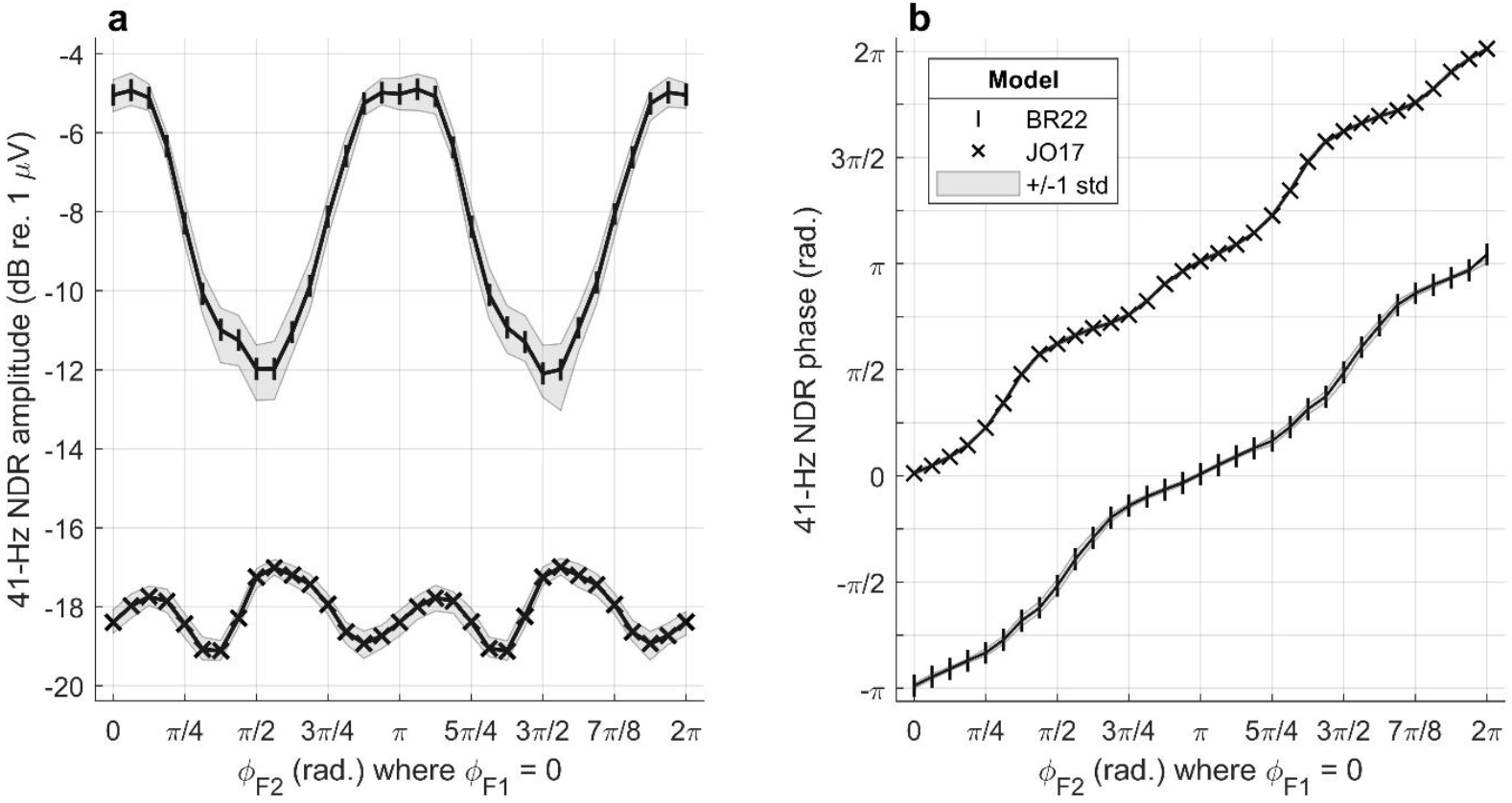
Modelling of the auditory nerve’s response to 60 seconds of ALFIES stimulation with the F1 starting AM phase fixed at ϕ_F1_ = 0 rad. and the F2 starting AM phase varied between ϕ_F2_ = 0 and 2π rad., the stimuli for which are depicted in Figure 5. Mean responses from the BR22 model are indicated with vertical symbols, whereas those from the JO17 model use crosses. Panel (a) shows the amplitude of the 41-Hz NDRs as a result of both models, whereas panel (b) shows the unwrapped phases of the 41-Hz NDRs. The grey shaded region depicts ±1 standard deviation from the mean in both panels. (NDR = neural distortion response, μV = microvolt, Hz = hertz, rad. = radian, std = standard deviation, ϕ = starting amplitude modulation phase)

### Experimental Methods

Nine cochlear-implant users took part, eight of whom were unilaterally implanted and one of whom was bilaterally implanted but for whom only one ear was assessed. A loudness-scaling procedure was conducted in the same manner as described in the General Experimental Methods to determine the current levels required for threshold and MCL for each of the pair of electrodes used. F1 and F2 AM rates were nominally set to 82 and 123 Hz, both amplitude modulated using 0.5*[1 + cos(2π *f_mod_* t + ϕ)] starting with ϕ= 0 such that the maximum current level for each electrode (MCL) was aligned temporally at the start of each 1-second epoch.

The neural origin of the NDR was first assessed by stimulating with F1 and F2 rates both at the nominal 82 and 123 Hz but also of 74 and 111 Hz, resulting in NDR frequencies of 41 and 37 Hz, respectively. Inclusion criteria required group delays significantly greater than 0 ms. Significance was confirmed on an individual participant level using bootstrapping as described in the General Experimental Methods.

For the subsequent conditions, the (*cosine*) starting AM phase of F1, ϕ_F1_, was fixed at 0 radians, whereas the starting AM phase of F2, ϕ_F2_, was varied between π/4, π/2, π, and 3π/4 radians. These F2 starting phases were selected based on the results of the modelling to include conditions that include minimum and maximum predicted NDR amplitudes and to include a whole cycle (from 0 to π radians) of variation. 300 epochs were recorded for each of these relative-phase conditions, unless significant responses were observed with narrow confidence intervals using the real-time analysis software in significantly fewer epochs (i.e. 150 epochs). When time allowed, additional relative-phase conditions were included, sometimes with as few as 90 epochs after observing significant responses, with ϕ_F2_ shifted to π/8, 3π/8, 5π/8, 7π/8, and/or 3π/2 radians.

An additional condition was included with a single 492-pps AM pulse train modulated at 41 Hz to capture the electrically-evoked auditory steady state response (eASSR) and compare its amplitude and phase to the distortion product responses elicited from the ALFIES stimuli. The artefacts were identified, blanked (50 µs prior and 900 µs after the onset of each stimulating pulse), and linearly interpolated in the time domain as described in detail in Arzounian *et al* [14]; the subsequent analysis pipeline involving fft and significance testing using Hotelling T^2^ were consistent with the analysis of the ALFIES stimuli.

### Experimental Results

Eight of the nine participants achieved the acceptance criterion described above, showing group delays greater than 0 ms for the NDRs and were retained in the subsequent analysis. The amplitudes of the eASSR and ALFIES NDRs can be observed in Figures 7a and 7b, respectively, and their phases in Figures 7c and 7d. Expect for two participants (C35R and C60R, ϕ_F2_ = 5π/8 and 7π/8), we were able to measure NDRs with F2 starting phases of ϕ_F2_ = 0 through π in increments of π/8. For three participants (C37R, C46R, and C72R) an F2 starting phase of ϕ_F2_ = 3π/2 was also included.

**Figure 7:**
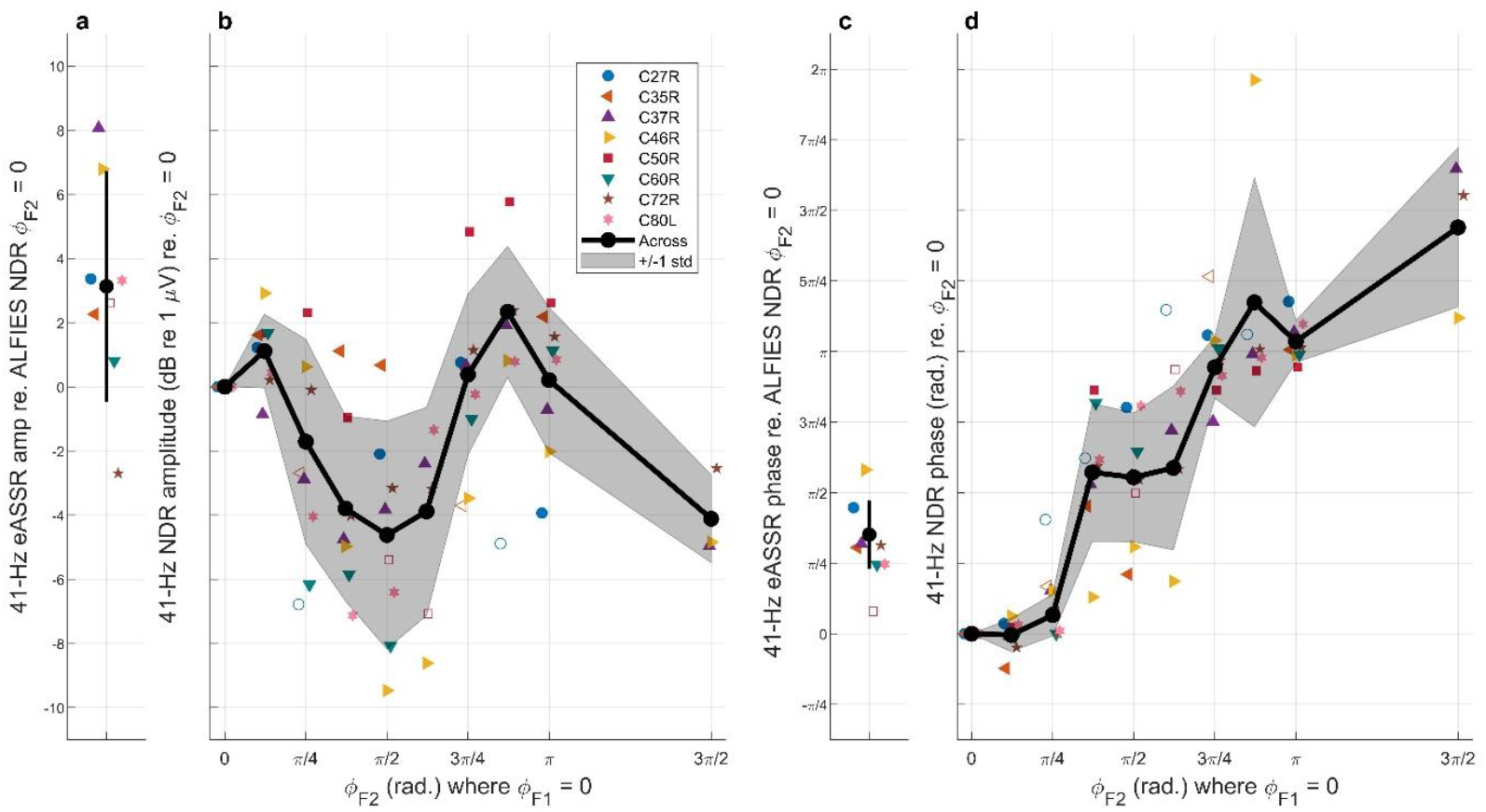
(a) Amplitudes of the eASSR at 41 Hz for individual participants in colors (throughout, open symbols represent observations that were not significant from the noise floor) and averaged across participants (black). Confidence intervals represent ±1 standard deviation across participants. (b) Amplitudes of the ALFIES NDR at 41 Hz for the conditions where the F1 pulse train was amplitude modulated with ϕ_F1_ = 0 whereas ϕ_F2_ varied between 0 and 3π/2. Data are plotted relative to the amplitude for the condition where ϕ_F2_ = 0. Black line represents the average across participants, and the shaded region represents ±1 standard deviation. (c) Phases of the eASSR at 41 Hz for individual participants (in colors) and averaged across participants (black). Confidence intervals represent ±1 standard deviation across participants. (d) Phases of the NDR at 41 Hz for the same conditions as depicted in panel B, plotted relative to the phase for the condition where ϕ_F2_ = 0. (NDR = neural distortion response, rad. = radian, std = standard deviation, ϕ_F1_ = F1 AM pulse train starting phase, ϕ_F2_ = F2 AM pulse train starting phase)

To compare the predictions of the two models with the experimental data, three LME models were fit to the NDR amplitudes (in dB re 1 μV) with participant specified as a random factor and an intercept fit for each. The *fitlme()* function in MATLAB version 2020b (Mathworks, Natick, MA, USA) was used with the default settings (i.e. the ‘FitMethod’ was set to ‘ML’). The first LME model fit two fixed effects: a cosine and sine curve with periods corresponding to one cycle per π radians of relative AM starting phase of the two interleaved pulse trains (cos2 and sin2, respectively). This sinusoidal behavior mimics the effect of relative phase estimated by using the BR22 model. The model structure was: y ~ sin2 + cos2 + (1 + sin2 + cos2 | subj), where y is the NDR amplitude in dB re 1 μV, and sin2 and cos2 are the sinusoid transformations of ϕ_F2_ with periods of π (sin2 = sin(2ϕ_F2_); cos2 = sin(2ϕ_F2_)). The model’s joint test of the two fixed effects was significant (F(2,60) = 12.9773, *p* < 0.0001), consistent with a significant modulation of the data with a period of π. The modulation amplitude was calculated as A = 2.414 dB re 1 μV (95% confidence intervals = [1.478, 3.527], calculated with 1000 bootstrapped samples with replacement), and starting phase as −0.132π radians (95% confidence intervals = [-0.296π, 0.0139π]). This suggests that the highest-amplitude NDRs are observed when the maxima of the AM pulse trains are aligned.

The second LME model fit two fixed effects: a cosine and sine curve with periods corresponding to *two* cycles per π radians of relative AM starting phase of the two interleaved pulse trains (cos4 and sin4, respectively). This sinusoidal behaviour mimics the effect of relative phase estimated by using the JO17 model. The model structure was: y ~ sin4 + cos4 + (1 + sin4 + cos4| subj), where sin4 and cos4 are sinusoids with periods of π/2 (sin4 = sin(4ϕ_F2_); cos4 = sin(4ϕ_F2_)). The model’s joint test of the two fixed effects (cos4 and sin4) was not significant (F(2,60) = 1.0355, *p* = 0.3613).

The third LME model directly fit the JO17 and BR22 model predictions as the two fixed factors to test directly whether each of the computational models predicted the behaviour or the NDR as observed in CI users. The model structure was: y ~ br22 + jo17 + (1 + br22 + jo17 | subj), where br22 contains the BR22 model predictions of the NDR amplitude as a function of relative AM starting phase, and jo17 contains the same model predictions from the JO17 model. An ANOVA leveraging the Satterthwaite method for degrees of freedom showed a significant effect of the BR22 model (F(1,8.383) = 17.242, *p* = 0.0029), with no significant effect of the JO17 model (F(1,56.44) = 0.0063, *p* = 0.937), nor the intercept (F(1,53.421) = 0.129, *p* = 0.721).

Taken together, the results from these three LMEs suggest that variations in NDR amplitude with relative phase are consistent with the predictions of the BR22 model, and provide no evidence that they are consistent with those of the JO17 model. Because refractoriness is dominant in the BR22 model while facilitation is dominant in the JO17 model, this suggests that refractoriness could be a stronger mechanistic contributor to the NDR than facilitation.

The amplitude and phase of the NDR when the maxima of the interleaved AM pulse trains was aligned was compared to the amplitude and phase of the eASSR responses using two-tailed t-tests across participants. For the amplitude this was not significant (t(6) = 2.297, *p* = 0.0613, α= 0.05/2 = 0.025), and the eASSRs and NDRs were equivalent within approximately 2.65 dB of each other (TOST (two-one-sided-t) test of equivalence within a 2.65-dB window: t(6) = ± 1.946, *p* = 0.0498). Hence, by judicious choice of relative AM phase the ALFIES method can produce an NDR with an amplitude comparable to that of the eASSR. The phase of the NDR and eASSRs differed significantly (t(6) = 7.658, *p* = 0.0003, α = 0.05/2 = 0.025), surviving corrections for two comparisons. Similar to Guérit *et al* [23], this phase difference was better predicted by the BR22 than the JO17 model.

## Experiment 3: The effect of relative phase on separated quadratic and cubic distortions

Experiment 3 investigated whether the effect of relative phase on the NDR amplitude could be explained by the linear summation of the separated QDR and CDR responses alone. The separated QDRs and CDRs were therefore recorded at 41 Hz and different relative phases in the same participants (except for C37R). These results could then be analytically combined to estimate the expected NDR amplitude given the amplitudes and phases of the separated distortion products as in equation (2):

,
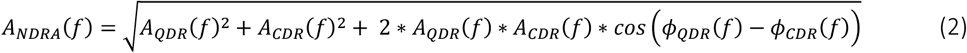

where *f* is the frequency of interest (in this case, 41 Hz), *A*_*QDR*_ and *A*_*CDR*_ are the amplitudes of the quadratic and cubic distortion products in the power spectrum, respectively, and *ϕ*_*QDR*_ and *ϕ*_*CDR*_ are their phases. This analytically combined NDR (*A*_*NDRA*_) can then be compared to the observed NDR (*A*_*NDRO*_) at the same frequency (41 Hz) recorded in experiment 2. This question was addressed first with the computational models and then investigated in CI users.

### Modelling Methods

Two frequency combinations were employed to achieve both QDRs and CDRs at 41 Hz. To observe the QDRs at 41 Hz, the F1 and F2 frequencies were set to 88 and 129 Hz, respectively, to achieve a QDR at 41 Hz and a CDR at 47 Hz. To observe the CDR at 41 Hz, the F1 and F2 frequencies were set to 76 and 111 Hz, respectively, to achieve a QDR of 35 Hz and a CDR of 41 Hz. This was done for the same relative AM phases as described in experiment 2, with the F1 modulation starting with ϕ_F1_ = 0 and the F2 modulation varying between ϕ_F2_ = 0 and ϕ_F2_ = 2π. For each relative-phase condition, the QDR and CDR at 41 Hz were then analytically combined according to equation 2.

### Modelling Results

The modelled amplitudes of the 41-Hz QDR and CDR components are depicted in Figure 8a as a function of ϕ_F2_, with the predictions of the BR22 and JO17 models shown by vertical line symbols (|) and crosses (x), respectively. Note that for both models, the QDRs remain relatively constant as ϕ_F2_ varies between 0 and 2π, whereas the CDRs do show some degree of variation, more pronounced in the JO17 model.

**Figure 8:**
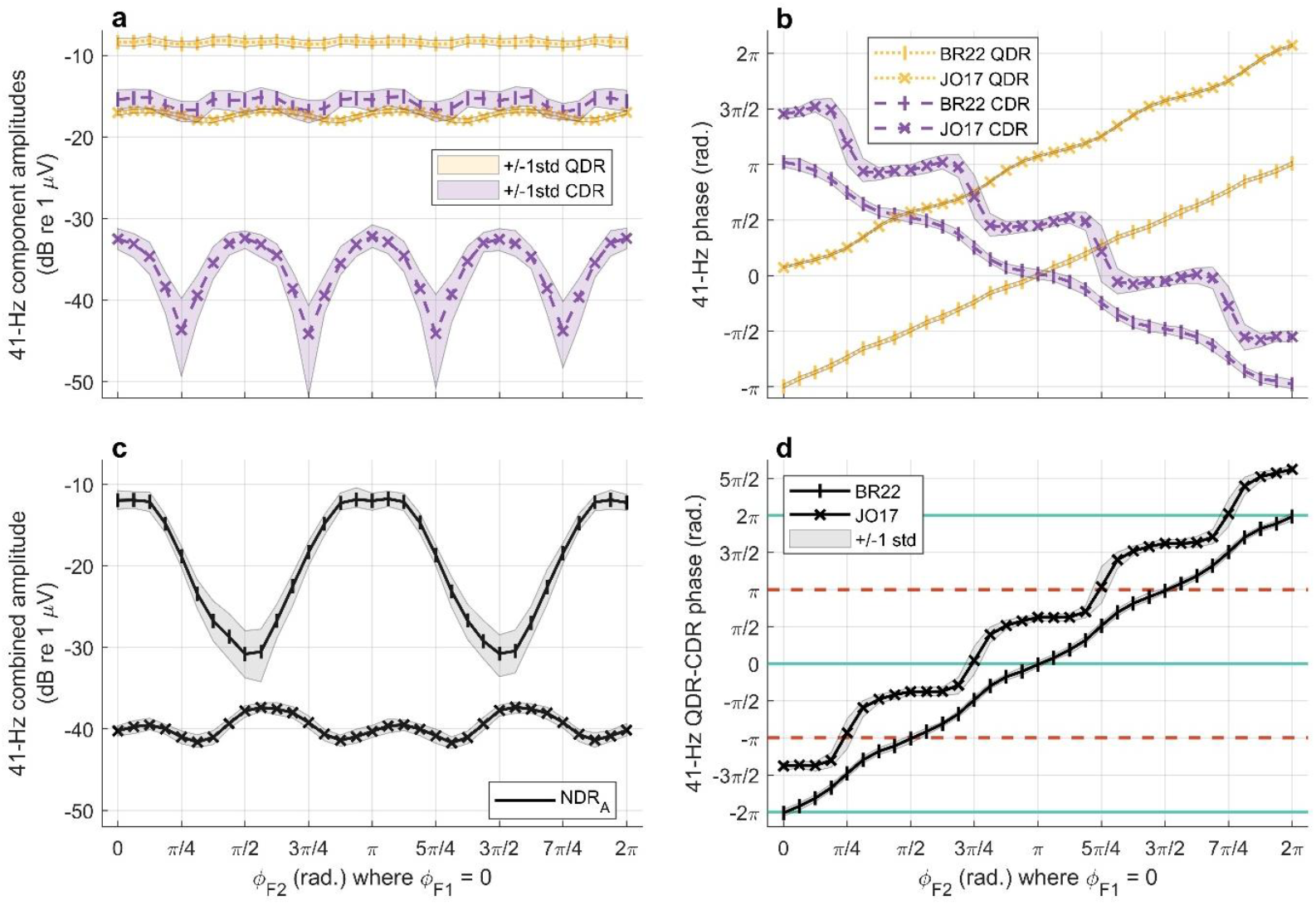
Modelling of the auditory nerve’s response to 60 seconds of ALFIES stimulation with the F1 starting AM phase fixed at ϕ_F1_ = 0 rad. and the F2 starting AM phase varied between ϕ_F2_ = 0 and 2π rad., with the 41-Hz QDR and CDR responses modelled separately and analytically combined. Responses from the BR22 model are indicated with vertical lines, whereas those from the JO17 model use crosses. Panel (a) shows the amplitude of the 41-Hz CDRs (purple, dashed lines) and QDRs (yellow, dotted lines) as a result of both models, with shaded regions indicating variation of ± 1 std across the 60 1-second simulated epochs. Panel (b) shows the unwrapped phases of the 41-Hz CDRs and QDRs from both models. Panel (c) shows the analytically-combined 41-Hz combined NDRs (black, solid line, NDR_A_) from both models when combined analytically from the separate CDR and QDR amplitudes and relative phases via equation (2), with shaded regions indicating variation of ± 1 std across the 60 1-second simulated epochs. Panel (d) shows the unwrapped phase differences (black, solid line) between the CDRs and QDRs from both models. Phase differences of 2π, 0, and −2 π indicate fully constructive interaction (solid teal lines),whereas those of –π and π indicate fully de-constructive interaction (dashed red lines). (std = standard deviation, NDR = neural distortion response, NDR_A_ = analytically-combined neural distortion response, CDR = cubic distortion response, QDR = quadratic distortion response, μV = microvolt, Hz = Hertz, rad. = radian)

Figure 8b shows the phases of both the 41-Hz QDR and CDR components as a function of ϕ_F2_. Both models predict that the QDR phase will increase a total of 2π as the ϕ_F2_ varies from 0 to 2π, although the two models predict QDR phases that are π offset from each other (BR22 starts at –π at ϕ_F2_ = 0 and JO17 starts at 0 at ϕ_F2_ = 0). In contrast, both models predict the CDR phases to *decrease* 2π over the course of the same ϕ_F2_ variation. The BR22 model predicts a relatively linear decrease starting at –π wherein the QDR and CDR align at ϕ_F2_ = 0, π, and again at 2π. The JO17 model predicts a non-linear decrease over the same ϕ_F2_ variation, with the QDR and CDR aligning at ϕ_F2_ = 3π/4 and 7π/4. To visualise the conditions wherein the QDR and CDR are in- and out-of-phase, Figure 8d plots the difference between the two phases for both models. Here it is clear to visualise that the BR22 model predicts the two distortion products to be in phase at ϕ_F2_ = 0, π, and 2π, and out of phase at ϕ_F2_ = π/2 and 3π/2. The JO17 model predicts them to be in phase at ϕ_F2_ = 3π/4 and 7π/4, and out of phase at ϕ_F2_ = π/4 and 5π/4.

As is formalised in equation (2), when added components are perfectly in phase with each other, they constructively interact, meaning that their amplitudes will sum. However, when added components are perfectly *out of* phase with each other, they interact destructively, meaning that their amplitudes will subtract. Using equation (2), the two separated CDR and QDR components were then analytically combined, and the result is displayed in Figure 8c. It appears that the variation in NDR amplitude calculated from summed QDR and CDR components fluctuate in a similar way for both models as in the combined NDR modelling displayed in Figure 6a, although the size of the fluctuations is amplified somewhat (from 5 to 18 dB) for the BR22 model. Importantly, as in Figure 6a, the variation in predicted NDR amplitude between 0 and π radians is repeated between π and 2π.

### Experimental Methods

The same participants that participated in experiment 2 also participated in experiment 3, with the exception of C37R. C36R was again excluded from analysis due to insufficient evidence that QDR and CDRs showed group delays greater than 0 ms. For all 7 retained participants, two F1 & F2 combinations were used to measure separated QDR and CDR of 41 Hz (same as for the modelling). Two to four relative-phase conditions were then included with F1’s modulation phase fixed at ϕ_F1_ = 0 at t = 0 ms, and F2 having a phase at t = 0 ms of ϕ_F2_ = 0 and ϕ_F2_ = π/2. When time allowed, additional conditions were included with ϕ_F2_ = π/4 (C60R, C46R, and C72R) and ϕ_F2_ = 3π/4 (C46R and C72R). Real-time analysis was employed as in previous experiments, and a range of durations was used for each condition from 90 to 300 1-second epochs.

The amplitudes of the NDR were then analytically combined from the phases and amplitudes of the separately-recorded QDRs and CDRs at 41 Hz to calculate *A*_*NDR*_ via equation 2. The resultant predicted phase of the combined QDRs and CDRs at 41 Hz were then also calculated via equation (3):

,
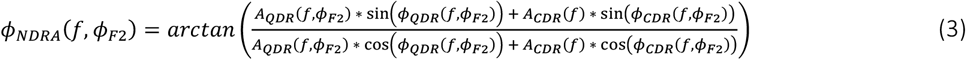

where, as in equation 2, *f* is the frequency of interest (in this case, 41 Hz), *A*_*QDR*_ and *A*_*CDR*_ are the amplitudes of the quadratic and cubic distortion product components in the power spectrum, respectively, and *ϕ*_*QDR*_ and *ϕ*_*CDR*_ are their phases. This is calculated as a function of *ϕ*_*F*2_, the starting phase of the pulse train amplitude-modulated at F2 Hz.

The *A*_*NDRA*_ and *ϕ*_*NDRA*_ were calculated for each ϕ_F2_ condition for each participant, regardless of significance of the QDR or CDR components. However, if either of these was insignificant, the analytically combined components were excluded from further analysis. Comparison between the analytical and observed amplitudes and phases were done only for the participants and ϕ_F2_ conditions where both were statistically significant from the noise floor. The primary comparisons of interest (investigated with LMEs) were whether there was evidence of *A*_*NDRA*_ differing substantially from *A*_*NDRO*_ or that *ϕ*_*NDRA*_ differed substantially from *ϕ*_*NDRO*_.

### Statistical Approach

To determine whether *A*_*NDRA*_ differed substantially from *A*_*NDRO*_ in the context of ϕ_F2_ variation, the significant NDR amplitudes were submitted to a Linear Mixed Effects (LME) model using the Statistical Package for Social Sciences (SPSS) Software version 28.0.0.0 (IBM, Armonk, New York, USA). *A*_*NDR*_ was set as the dependent variable, ϕ_F2_ starting phase was set as a fixed effect with 4 levels (ϕ_F2_ = 0, π/4, π/2, and 3π/4), observation type was set as a fixed effect with 2 levels (analytically combined vs observed), and an interaction between ϕ_F2_ starting phase and observation type was included, as was an intercept, and participant was specified as a random factor. Post-hoc pairwise comparisons were corrected for multiple comparisons using the Bonferroni-Dunn method. The same model was fit for *ϕ*_*NDR*_ set as the dependent variable. A significant interaction would provide evidence that variation in *A*_*NDR*_ as a function of ϕ_F2_ *cannot* be explained solely by the linear summation of the quadratic and cubic distortions that would be primarily driven by the relative phases of the two components that determines the degree of constructive vs deconstructive interaction.

### Experimental Results

For two participants (C27R and C35R), no significant CDR components were observed either at ϕ_F2_ = 0 or ϕ_F2_ = π/2 radians. Additionally, no significant combined NDR component was observed at 41 Hz for the ϕ_F2_ = π/2 radians condition for C50R. Note that this reduced the sample size of conditions where both *A*_*NDRA*_ and *A*_*NDRO*_ were significant to N=5 for ϕ_F2_ = 0, to N=3 for ϕ_F2_ = π/4, to N=4 for ϕ_F2_ = π/2, and to N=2 for ϕ_F2_ = 3π/4.

The separately recorded QDR and CDR amplitudes and phases as a function of ϕ_F2_ are depicted in Figures 9a and 9b, respectively. Figure 9c depicts the *A*_*NDRA*_ values for each participant calculated via equation 2, as well as the observed *A*_*NDRO*_ values that are also displayed in Figure 7b. Figure 9d depicts *ϕ*_*NDRA*_ values for each participant calculated via equation 3, as well as the observed *ϕ*_*NDRO*_ values that are also displayed in Figure 7d. In both cases the predicted amplitudes and phases broadly follow those obtained. This observation is reflected by a lack of a significant interaction between measure (obtained vs predicted) and ϕ_F2_, either for the LME with *A*_*NDR*_ as the dependent variable (F(3,30) = 0.58, *p* = 0.63) or for the one with the *ϕ*_*NDR*_ as the dependent variable (F(3,30) = 1.76, *p* = 0.18). The main effect of measure (obtained vs predicted) was also not significant in either case (*A*_*NDR*_ F(1,30)=2.58, p=0.12; *ϕ*_*NDR*_ F(1,30)=0.044, p=0.84). Therefore, at the group level the amplitudes and phases of the NDR produced when the QDR and CDR overlap are consistent with those components summing linearly, according to equations (2) and (3), with the caveat of the small sample size and notable exceptions (i.e. C46 ϕ_F2_ = π/4, C60R ϕ_F2_ = 0, and C72R ϕ_F2_ = π/4 and 3π/4).

**Figure 9:**
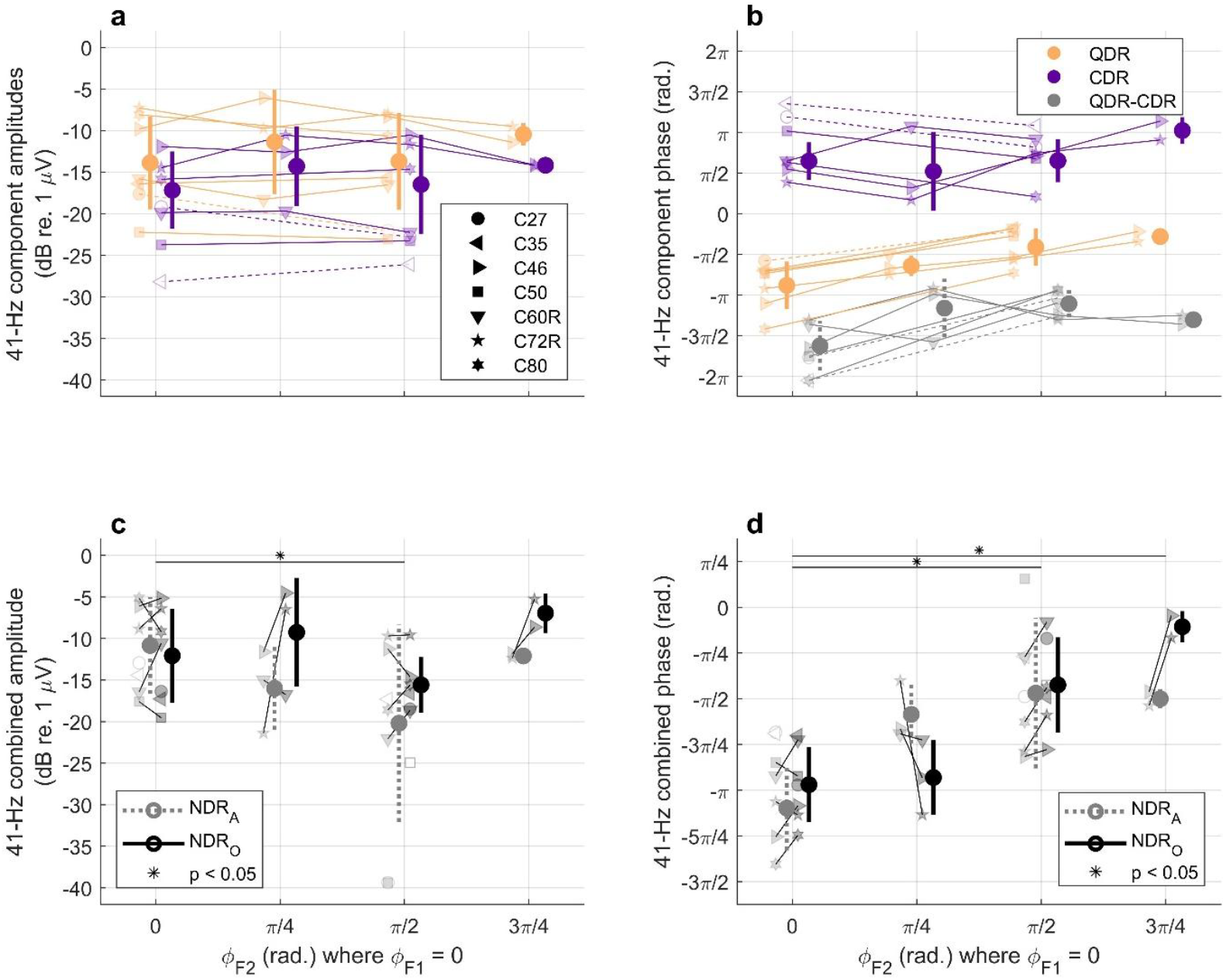
(a) separately recorded 41-Hz QDR (yellow) and CDR (purple) response amplitudes (dB re 1 μV) as a function of ϕ_F2_ values (x-axis). Across-participant means are plotted with large solid circles, where error bars represent ± 1 standard deviation (throughout all panels). Each of the transparent symbols represents an individual participant (key used throughout all panels), with light, narrow lines separating the observations from the same participants across ϕ_F2_ values. (b) The phases of the separately recorded 41-Hz QDR (yellow) and CDR (purple) components, as well as their phase difference (grey, dotted lines) analogous to the modelling predictions in Figure 8d. (c) Composite 41-Hz NDR amplitudes (dB re 1 μV) analytically combined from the data in panels A and B via equation (2) and analogous to the modelling in Figure 8c (NDR_A_, grey, dotted lines), and observed when quadratic and cubic distortions occur at the same frequency, analogous to the modelling in Figure 6a (NDR_O_, black, solid lines). Asterisks indicate significant differences. (d) Composite 41-Hz NDR phases (rad.) analytically combined from data in panels A and B via equation (3) (NDR_A_, grey, dotted lines), and observed when quadratic and cubic distortions occur at the same frequency (NDR_O_, black, solid lines). Asterisks indicate significant differences. (dB = decibels, rad. = radian, NDR_A_ = analytically combined neural distortion response, NDR_O_ = observed neural distortion response, ϕ_F2_ = starting amplitude-modulation phase of the F2 pulse train, ϕ_F2_ = starting amplitude-modulation phase of the F1 pulse train, * p < 0.05)

## Experiment 4: The effect of alternating polarity

The objective of the fourth and final experiment was to assess the effect on the NDR of alternating the leading polarity of the two interleaved, AM pulse trains. In all of the experiments reported so far the biphasic F1 and F2 pulses were always in cathodic-leading polarity, with the result that for each pulse pair the adjacent phases of the two pulses had opposite polarity. Here we compare this “fixed polarity” stimulation to an “alternating polarity” condition where the second pulse in each pair was anodic-leading, potentially producing summation between the two central anodic phases. Evidence for this increased facilitation or summation comes from a psychophysical experiment by Macherey *et al* who reported substantially lower MCLs for trains of pulse pairs where the central phases were anodic compared to when one was anodic and the other cathodic [29].

The hypothesis was that for the same stimulating current levels, the constructive between-pulse interaction in this second ‘alternating-polarity’ condition would result in a larger NDR amplitude than in the control condition where both stimulating pulses had a cathodic-leading polarity. Another model-based hypothesis was that if the alternating polarity leads to a large amount of facilitation, such that this mechanism comes to dominate the between-pulse interaction between F1 and F2 components, the phase of the NDR response may change compared to the same-polarity condition. This is because the models predicted that if facilitation dominates the NDR generation (as in JO17) in the alternating-polarity condition, the NDR phase should be shifted by π compared to the fixed-polarity condition where refractoriness dominates NDR generation (as in BR22, see Figure 6b).

### Modelling Methods

For this experiment, two conditions were assessed with the JO17 model. Both used a stimulus onset asynchrony (SOA) of 100 μs instead of the 200 μs used in experiments 1-3. This was shortened to reduce the time between stimulating pulse phases in the F1-F2 pulse pairs to a range where the responsive neurons may more-effectively integrate the charge of temporally adjacent pulses. The control “fixed polarity” (FP) condition used biphasic stimulating pulses using an anodic leading polarity, whereas in the ‘alternating-polarity’ (AP) condition the second (F2) biphasic pulses’ leading polarities were set to cathodic and the resulting F1-F2 pulse-pairs contained a central cathodic phase. This was done because the JO17 model is fit using parameters determined from deafened animals, where cathodic stimulation is more effective than anodic stimulation [30], opposite to what is commonly seen in humans [31]. Phase durations of 25 μs and inter-phase-gaps of 8 μs were used, resulting in an inter-pulse interval (IPI) of 42 μs. The BR22 model was not included because it does not contain polarity sensitivity and therefore would not be able to assess the effect of this manipulation.

As in experiments 2 and 4, the JO17 model was run for 2 seconds (with the 1^st^ onset second discarded from analysis) for 60 repetitions, to allow for characterisation of variability.

### Experimental Methods

For the experimental stimulation, a control condition was included with all biphasic stimulating pulses using a *cathodic* leading polarity, as in all previous experiments. A second condition was included with the leading polarity of the F2 pulse (the second of the pair) switched to the anodic phase, resulting in a central anodic phase for the pairs of F1-F2 pulses. SOA was 100 µs and phase durations varied as described in Table 1, therefore inter-pulse intervals (IPIs) between the pairs of pulses of the AM pulse trains (the time between the offset of the first pulse and the onset of the second) varied between 8 (i.e. C46) and 42 (i.e. C60) μs, with the majority (6 of 8) of participants using 18 μs IPIs (see Table 1 and Figure 10 for details).

**Figure 10:**
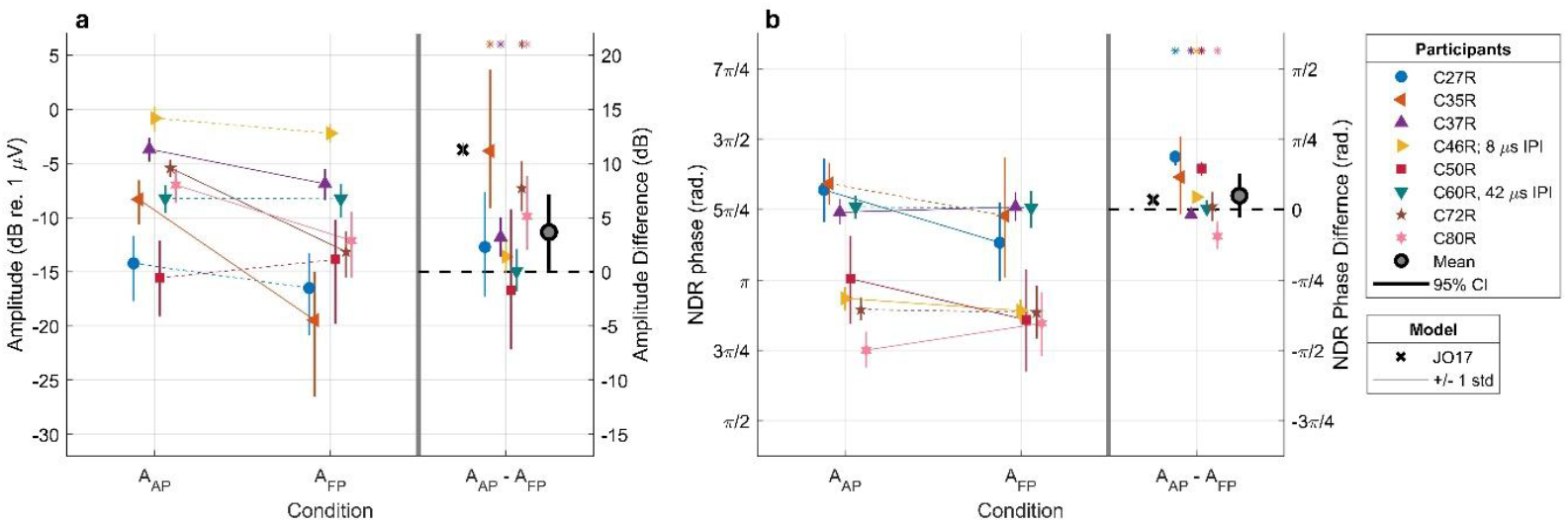
(a) A_AP_ and A_FP_ amplitudes (in dB re 1 μV) for 41-Hz NDRs with 100 μs SOAs (left), as well as their pairwise differences (right). (b) ϕ_AP_ and ϕ_FP_ phases (in radians) with 100 μs SOAs (left), as well as their pairwise differences. Throughout, each coloured symbol represents an individual participant, and associated error bars indicate the central 95% of their bootstrapped distribution. The thick black circles with grey faces indicate the across-participant pairwise difference means, and the thick black bars indicate across-participant 95% confidence intervals. Note that after corrections for multiple comparisons, neither the across-participant amplitude nor phase difference between the alternating-polarity and control conditions was significantly greater than 0 dB or radians, respectively. Black crosses indicate the modelling predictions from the JO17 model, with grey lines indicating ± 1 std across 60 repetitions. When not specified (for 6 of 8 participants), IPIs were 18 μs. (dB = decibels, μV = microvolts, rad. = radians, 95% CI = 95% confidence intervals, ϕ = phase, A = amplitude, IPI = inter-pulse interval, std = standard deviation, * indicates that the central 95% of the bootstrapped distribution excludes 0)

The same 9 CI users that participated in experiment 2 took part, with the same inclusion criteria. The participants underwent the same loudness-scaling procedure as described before but with the alternating-polarity stimuli, and the MCLs so derived were used for both the AP and fixed-polarity (FP) conditions. All stimulation was done with an F1 and F2 AM frequency of 82 and 123 Hz, respectively, and both pulse trains were amplitude modulated with 0.5*[1 + cos(2π *f_mod_* t + ϕ)] with a starting phase of ϕ = 0π. This meant that the NDR frequency was 41 Hz and the amplitude modulations of F1 and F2 had their peaks aligned (as in Figure 5a).

The amplitude of the 41-Hz component from the FP condition’s power spectrum (*A*_*FP*_) was subtracted from that of the AP condition (*A*_*AP*_). This was done both on an individual participant basis employing bootstrapping, as well as across participants for the entire full-epoch datasets for the two conditions.

## Results

Of the nine participants who completed the experiment, eight met the inclusion criteria and were included in subsequent analysis. Some participants spontaneously reported softer perceived loudness for the standard fixed-polarity condition. Note that for these two conditions, the same stimulating current levels were used for both the FP and AP stimuli. However, these perceived-loudness reports were not methodologically recorded, nor requested from every participant.

Figure 10a shows the *A*_*AP*_ and *A*_*FP*_ amplitudes, as well as their pairwise differences. Figure 10b likewise shows the *ϕ*_*AP*_ and *ϕ*_*FP*_ phases and their pairwise differences. These are shown for individual participants, where the confidence intervals are calculated via 1000 repetitions of bootstrapping, as well as across all eight participants retained in the analysis.

A two-tailed t-test for the difference between NDR amplitudes (*A*_*AP*_ - *A*_*FP*_) across participants was not appropriate due to evidence of deviation from a normal distribution using a Kolmogorov-Smirnov normality test (k-stat = 0.67, critical value = 0.45, *p* = 0.0005). A non-parametric Wilcoxon signed rank test on the difference between NDR amplitudes (*A*_*AP*_ - *A*_*FP*_) across participants was performed instead, which found a difference significantly higher than 0 dB (signed rank = 33, *p* = 0.0391). This result does not survive corrections for multiple comparisons (as we also tested for phase differences) where the significance threshold is shifted to α = 2/0.025. The average size of the difference was about 3.65 ± 3.51 dB (variation representative of 95% confidence intervals calculated *without* correction for multiple comparisons).

A Komogorov-Smirnov normality test did not demonstrate evidence that the phases of these responses (*ϕ*_*AP*_ - *ϕ*_*FP*_) differed from a normal distribution (k-stat = 0.38, critical value = 0.45, *p* = 0.15). Therefore a standard two-tailed t-test was performed, which did not reveal a difference between the conditions (t(7) = 1.47, 95% confidence intervals = −0.93 – 0.40 radians, *p* = 0.184), contrary to our hypothesis of a change in dominance of the underlying mechanism (from refractoriness to facilitation).

## Discussion

### Summary

We explored mechanisms underlying the generation of neural distortions (NDRs) to stimulation with alternating-frequency, interleaved, amplitude-modulated pulse trains (“ALFIES” stimuli) in cochlear implant users. The results provide insight into the temporal integrations at play, in addition to highlighting stimulation parameters important for optimising the SNR of NDRs.

The first experiment demonstrated that both quadratic and cubic neural-distortion responses can be observed using the ALFIES technique. Previous work had not isolated multiple distortion products [9], [11], whereas here the F1 frequency was shifted from 80 to 82 to isolate QDRs (38 Hz) and CDRs (44 Hz) from each other. Quadratic distortions were generally larger and more frequently observed than cubic distortions.

The observation of a relatively consistent phase difference between the QDR and CDRs (and therefore of possible cancellation between the two) inspired the second experiment. That revealed an effect of relative AM phase between the F1- and F2-AM modulated pulse trains on the amplitude of the NDR (when QDR and CDR are constrained to the same frequency). At the group level, the highest SNRs were observed when the maxima of the AM were aligned between the two interleaved pulse trains. A comparison of the data to the predictions of two computational models provided evidence that refractoriness (but not facilitation) was likely responsible for the interaction between F1 and F2 pulse trains that was necessary for NDR generation. Experiment 2 also showed that optimal selection of relative-AM phase can give rise to ALFIES NDR amplitudes comparable to those observed from eASSRs with the same stimulation parameters (within a 2.65 dB window).

The third experiment investigated interaction effects between separated distortion products and relative AM phase. It showed that, at the group level, the NDRs when the CDR and QDR fell at the frequency were consistent with a simple sum of the two components. However this conclusion was tempered by the small sample size and idiosyncratic effects of relative phase in some participants. The fourth and final experiment investigated the effects of alternating the leading polarity between the F1 and F2 pulses to create a centrally effective phase. This manipulation produced idiosyncratic effects across participants, resulting in increases ranging between 3.2 and 11.1 dB in the NDR amplitude relative to the fixed-phase condition in 4 participants, with no effect in the remaining 5; this effect did not survive correction for multiple comparisons at the group level.

### Limitations and theoretical implications

The experimental and modelling results presented here indicate that refractoriness could account both for the interaction between F1 and F2 pulses and the nonlinearity that are necessary for the generation of an NDR. If this is correct, it shows that the NDR can arise from a single process in which the temporal dependency and nonlinearity are not separate processes, as originally assumed, but intertwined. In the context of the BR22 model, this arises from spike generation which is inherently nonlinear (incorporating rectification, as there are no ‘negative spikes’) on one hand, and leads to refractoriness (a temporal dependency) on the other. However, it is not possible to rule out alternative or additional mechanisms. For example, at the level of the auditory nerve the effects of accommodation may be qualitatively similar to those of refractoriness, in that the response to a pulse may be reduced by the presentation of a preceding, high-amplitude pulse. However, evidence against this explanation comes from findings that accommodation – at least in animal studies – increases with increasing inter-pulse interval up to about 500 μs, whereas the NDR is constant for IPIs between 8 and about 200-400 us, after which it decreases [11]. In addition, the fact that scalp-based EEG recordings are less sensitive to auditory-nerve and brainstem responses than to cortical responses means that we are necessarily recording the cumulative effects of multiple processing stages, any or all of which may be highly nonlinear. What we *can* conclude, however, is that the temporal dependencies necessary for the NDR generation are likely present at the level of the auditory nerve, and that the spike generation and refractoriness known to operate at that level are sufficient to explain the pattern of results described here and previously. Future experiments either with implanted recording electrodes in animals or perhaps tympanic-membrane electrodes with humans may be able to more selectively measure the effects of peripheral processes in isolation.

A second consideration is that the present results were obtained at a single pulse rate and (except for experiment 4) a single IPI of 200 μs. It is therefore possible that the relative roles of refractoriness and facilitation would be different at either shorter or longer IPIs. However, a re-analysis of the data reported by Guérit et al [11] revealed that the phase of the NDR was approximately constant for IPIs ranging from 8 to 948 μs (not shown here). As argued due to the modelling results from Experiment 2, a change in dominant mechanism between refractoriness and facilitation would lead to a phase shift, and this was not observed here.

### Practical and clinical implications

Both the eASSR and ALFIES have been proposed as objective measures of loudness [11], [14], [32], with the potential clinical application of informing the audiological fitting of MCLs in patients unable to make reliable subjective responses. For example, the eASSR has been shown to correlate with acoustic ASSRs and subjective loudness judgements in patients with a CI in one ear and normal hearing in the other [33], and the slope of the eASSR amplitude growth function correlates with the subjective dynamic range in long-term CI users [14]. However the eASSR requires high-rate sampling systems to isolate the neural response from the CI electrical artefacts, is limited to moderate carrier pulse rates, and produces artefacts that are larger when recorded ipsilateral compared to contralateral to the CI. ALFIES overcomes these problems but, in previous studies, produced an NDR that had a lower amplitude than the eASSR. The present study shows that a judicious choice of relative phase increases the NDR amplitude so as to be comparable to the eASSR, while largely avoiding the electrical artefact and allowing measurement using a technique that permits wide range of pulse rates and recording montages.

## Data Availability

Data will be made available upon request within institutional restrictions. Computational Modelling Code Associated with this manuscript can be found on Zenodo.

https://doi.org/10.5281/zenodo.23695046

## Acknowledgements

We thank Robin Gransier, Jan Wouters, Matthew Richardson, and John Middlebrooks for helpful discussions. We also thank the cochlear-implant users who volunteered their time to participate in this research.

## Author Contribution

CG: Conceptualization, Data curation, Formal analysis, Software, Methodology, Investigation, Visualization, Project administration, Writing – original draft, Writing – review & editing, FG: Conceptualization, Formal analysis, Writing – review & editing, DA: Investigation, Supervision, Writing – review & editing, RPC: Conceptualization, Resources, Supervision, Funding acquisition, Writing – review & editing

## Data Availability

Data will be made available as possible upon request, within institutional and ethical board restrictions.

## Code Availability

Code for the modelling work presented in this manuscript is available [28].

## Artificial Intelligence

No artificial intelligence was used in the preparation of this manuscript or in any of the work presented within the manuscript.

